# Epigenomic signature of major congenital heart defects in newborns with Down syndrome

**DOI:** 10.1101/2023.05.02.23289417

**Authors:** Julia S. Mouat, Shaobo Li, Swe Swe Myint, Benjamin I. Laufer, Philip J. Lupo, Jeremy M. Schraw, John P. Woodhouse, Adam J. de Smith, Janine M. LaSalle

**Author notes:** Corresponding author: Janine LaSalle. Equal contributors (Adam J. de Smith, Janine M. LaSalle).

## Abstract

**Background:** Congenital heart defects **(CHDs)** affect approximately half of individuals with Down syndrome **(DS)** but the molecular reasons for incomplete penetrance are unknown. Previous studies have largely focused on identifying genetic risk factors associated with CHDs in individuals with DS, but comprehensive studies of the contribution of epigenetic marks are lacking. We aimed to identify and characterize DNA methylation differences from newborn dried blood spots **(NDBS)** of DS individuals with major CHDs compared to DS individuals without CHDs.

**Methods:** We used the Illumina EPIC array and whole-genome bisulfite sequencing **(WGBS)** to quantitate DNA methylation for 86 NDBS samples from the California Biobank Program: 1) 45 DS-CHD (27 female, 18 male) and 2) 41 DS non-CHD (27 female, 14 male). We analyzed global CpG methylation and identified differentially methylated regions **(DMRs)** in DS-CHD vs DS non-CHD comparisons (both sex-combined and sex-stratified) corrected for sex, age of blood collection, and cell type proportions. CHD DMRs were analyzed for enrichment in CpG and genic contexts, chromatin states, and histone modifications by genomic coordinates and for gene ontology enrichment by gene mapping. DMRs were also tested in a replication dataset and compared to methylation levels in DS vs typical development **(TD)** WGBS NDBS samples.

**Results:** We found global CpG hypomethylation in DS-CHD males compared to DS non-CHD males, which was attributable to elevated levels of nucleated red blood cells and not seen in females. At a regional level, we identified 58, 341, and 3,938 CHD-associated DMRs in the Sex Combined, Females Only, and Males Only groups, respectively, and used machine learning algorithms to select 19 Males Only loci that could distinguish CHD from non-CHD. DMRs in all comparisons were enriched for gene exons, CpG islands, and bivalent chromatin and mapped to genes enriched for terms related to cardiac and immune functions. Lastly, a greater percentage of CHD-associated DMRs than background regions were differentially methylated in DS vs TD samples.

**Conclusions:** A sex-specific signature of DNA methylation was detected in NDBS of DS-CHD compared to DS non-CHD individuals. This supports the hypothesis that epigenetics can reflect the variability of phenotypes in DS, particularly CHDs.

## 1. Introduction

Down syndrome **(DS)** is a set of distinct clinical features that result from trisomy 21, the most common autosomal aneuploidy across live births. Clinical characteristics of DS vary across individuals but include intellectual disability, short stature, muscle hypotonia, atlantoaxial instability, reduced neuronal density, cerebellar hypoplasia, and congenital heart defects **(CHDs)** (1). CHDs affect ∼50% of newborns of both sexes with DS (2–5) despite their diagnosis in only ∼1% of newborns without DS (6). The most frequently diagnosed CHD in children with DS is an atrioventricular septal defect **(AVSD)**, a condition characterized by a large hole in the heart due to improper development of the endocardial cushion. Many cases of DS-CHDs, particularly AVSD, are diagnosed *in utero* by ultrasound, but others are not diagnosed until after birth following obvious symptoms or an echocardiogram and often require surgery.

The mechanisms influencing the development of CHDs among individuals with DS are not clear. Studies of partial trisomy 21 patients have pinpointed critical regions on chromosome 21, including the Down syndrome cell adhesion molecule (*DSCAM*) gene, that appear to underlie CHD development (7), but these have not addressed the incomplete penetrance among individuals with complete trisomy 21. Additionally, genome-wide association studies and candidate-gene approaches have identified variants on chromosomes throughout the genome that are associated with CHDs in DS (8–13). However, these genetic variants do not sufficiently explain CHD risk among those with DS.

Another molecular driver or biomarker of CHD risk in children with DS may be epigenetic mechanisms such as DNA methylation. Increasing evidence has shown epigenetic alterations and gene-environment interactions to be involved in the pathogenesis of non-syndromic CHDs (14,15), but comprehensive studies of genome-wide DNA methylation variation associated with DS-CHD are lacking. We previously used whole-genome bisulfite sequencing **(WGBS)** of newborn dried blood spots **(NDBS)** to examine methylation profiles in 11 DS-CHD compared to 10 DS non-CHD samples, as part of a larger DS vs typical development **(TD)** study (16). There were 1,588 nominally significant (*p* <0.05) differentially methylated regions **(DMRs)** (35% hypermethylated, 65% hypomethylated) distinguishing DS-CHD from DS non-CHD and these regions were enriched for terms related to the heart, as well as neurodevelopment and metabolism (16). These promising but preliminary results suggesting an epigenomic signature of CHD within DS led us to conduct the present study.

This current study used WGBS of NDBS obtained from the California Biobank Program among 86 DS individuals with and without major CHDs to identify specific loci, biological pathways, and genic contexts that are associated with risk for CHDs in the DS population. Very few studies have conducted WGBS on NDBS, a sample source that is accessible, widely banked, reflective of the intrauterine period, and informative regarding dysregulation in other tissues, including the brain and the heart (16). In contrast to reduced representation methods such as arrays, this WGBS study provides insight to the entire DS-CHD epigenome, particularly because regional methylation smoothing approaches increase confidence over regions with relatively low coverage. Additionally, our study investigates similarities/differences in molecular signatures of DS-CHD in males compared to females, as well as DS-CHD compared to DS (versus TD). Our findings showed sex-specific global and region-specific changes to methylation that may serve as biomarkers and/or be functionally important in the development of CHDs in individuals with DS.

## 2. Methods

### 2.1 Study Populations and DNA extraction from NDBS

This study was approved by Institutional Review Boards at the California Health and Human Services Agency, University of Southern California, and University of California, Davis. For the Discovery study, deidentified NDBS were obtained from 90 newborns with DS from the California Biobank Program (CBP, SIS request number 572), with a waiver of consent from the Committee for the Protection of Human Subjects of the State of California (17). Demographic and birth-related data, including sex, race/ethnicity, birthweight, gestational age, and age of blood collection were obtained from the CBP **(Supplemental Tables S1-S2)**. DS newborns with CHD or without CHD were identified via linkage between the California Department of Public Health Genetic Disease Screening Program and the California Birth Defects Monitoring Program (CBDMP). In brief, the CBDMP is a population-based surveillance program that covers ∼30% of the births in California, including 10 counties, which are representative of the state’s population (18). Birth defects diagnosis data from CBDMP for the 90 newborns were coded into “major birth defects” and “major heart defects” using guidelines from the National Birth Defects Prevention Network (6). Major defects included AVSD and tetralogy of Fallot. We identified 46/90 newborns with a CHD, of which 44 were AVSDs, and 3 had tetralogy of Fallot. For this study, we focused on major heart defects and following sample QC (described below) we included 45 DS with CHD (27 female, 18 male) and 41 DS without CHD (27 female, 14 male). DNA was extracted from one 4.7 mm card punch of each of the 90 NDBS, roughly 1.4cm in diameter, with the Beckman Coulter GenFind V3 Reagent Kit (cat #C34880).

### 2.2 Whole genome bisulfite sequencing

All DNA samples were sonicated to ∼350bp with a peak power of 175, duty of 10%, 200 cycles/burst, and a time of 47 seconds. The sonicated DNA was cleaned and concentrated with Zymo gDNA clean and concentrator columns and eluted in 25 μl EB. Bisulfite conversion was performed with the Zymo EZ DNA Methylation Lightning Kit (cat #11-338) using ∼35 ng of each sonicated sample. Libraries were prepared using the Swift ACCEL-NHS MethylSeq DNA Library Kit (cat #30096) with 7 cycles of PCR for normal-input samples and 11 cycles for low-input samples. Libraries were pooled and a 0.85X SPRI cleanup was performed on 250 μl of the pool, eluted in 100 μl. The library pool (concentration of 3.63 ng/ μl) was sequenced across 4 lanes of an Illumina NovaSeq 6000 S4 flow cell using 150 bp paired end reads.

FASTQ files for each sample were merged across lanes using FASTQ_Me (19) and aligned to the hg38 genome using CpG_Me (20) with the default parameters (21–24). The alignment pipeline includes trimming adapters and correcting for methylation bias, screening for contaminating genomes, aligning to the reference genome, removing PCR duplicates, calculating coverage and insert size, and extracting CpG methylation to generate a cytosine report (CpG count matrix) and a quality control **(QC)** report. Global methylation for each sample was calculated as the total number of methylated CpG counts divided by the total number of CpG counts from CpG count matrices. From the 90 samples sequenced, four samples were removed from analysis: two due to high levels of sequence duplication and two due to missing sample data.

### 2.3 Genome-wide DNA methylation arrays

In addition to WGBS, existing DNA methylation data was available from NDBS for each sample from Illumina Infinium MethylationEPIC (EPIC) DNA methylation arrays (17). In brief, DNA was isolated from a separate one-third portion of the NDBS, bisulfite conversion performed as above, and DNA samples were block-randomized (ensuring equivalent distribution of sex and race/ethnicity on each plate) for EPIC arrays (17). QC of DNA methylation array data was conducted in R using “minfi”, “SeSAMe”, and “noob” packages, and trisomy 21 was confirmed from copy-number variation plots generated using the “conumee” package, as described (17). DMRs associated with DS-CHDs were investigated using the ipDMR method with the ENmix R package (25).

### 2.4 Cell type estimation

To estimate nucleated cell proportions in NDBS samples, we used the EPIC array data to perform reference-based deconvolution using the Identifying Optimal Libraries (IDOL) algorithm (26). Briefly, “estimateCellCounts2” function from the “FlowSorted.Blood.EPIC” R package was used to estimate proportions of CD8+ T lymphocytes **(CD8T)**, CD4+ T lymphocytes **(CD4T)**, natural killer **(NK)** cells, B lymphocytes **(B cell)**, monocytes, granulocytes, and nucleated red blood cells **(nRBC)**, using cord blood cell reference samples included in the “FlowSorted.CordBloodCombined.450k” R package.

### 2.5 Sample trait analysis

Newborn sample traits of global CpG methylation, birthweight, gestational age of delivery, age of blood collection, race, ethnicity, and cell type proportions were correlated using Pearson’s method with the Hmisc package v4.7.1 and *p*-values were adjusted by FDR (0.05 threshold) using the corr.test function in the Psych package v2.2.9 in Rv4.1.3. DS-CHD vs DS non-CHD samples (sex-combined and sex-segregated) were tested for differences across sample traits using Welch’s unpaired variances *t*-test with GraphPad Prism v9.4.1. Stepwise forward logistic regression was performed to determine the variables that best predicted CHD in each sex using the glm (family = binomial) function in R v4.1.3. Stepwise linear regression was performed to determine the variables that best predicted global CpG methylation in each sex using the lm function in R v4.1.3.

### 2.6 DMR analysis from WGBS

DMRs for DS-CHD vs DS non-CHD in the WGBS data were called for Sex Combined, Females Only, and Males Only samples using DMRichR v1.7.3 (16) and R version 4.1.0. Default parameters were used to identify DMRs containing at least 5 CpGs with at least a 5% methylation difference between groups, with each CpG requiring at least 1x coverage in at least 75% of samples. DMRichR uses bsseq (27) to extract methylation levels from cytosine reports and dmrseq (28) to identify DMRs. The dmrseq algorithm detects candidate regions whose smoothed pooled methylation proportion show differences between groups, then assesses the significance of candidate regions through permutation testing of the pooled null distribution to calculate p-values that are then FDR corrected to generate q-values (28). In all three comparisons (Sex Combined, Females Only, and Males Only), we adjusted for sample traits that were correlated with global methylation (|r| >0.2): age of blood collection and all cell types. Sex was additionally adjusted for in the Sex Combined analysis. Gestational age and birthweight met this cut-off in males, but not females, and were not corrected for because the effect of gestational age on DNA methylation has been found to mostly be due to nRBC proportion (29) and birthweight is largely dependent on gestational age. Sex chromosomes were included in Females Only and Males Only comparison but not the Sex Combined comparison. The sex of each sample was confirmed by the number of reads of sex chromosomes as previously described (16).

Principal component analysis **(PCA)** was performed using smoothed methylation values over the DMRs identified in each comparison to test for separation of CHD and non-CHD samples. Data was standardized so each variable had a mean of 0 and standard deviation of 1, and principal components were selected by parallel analysis from 1000 permutations using GraphPad Prism v9.4.1. The two principal components that explained the greatest variance in the data were selected for graphing and samples were color-coded by CHD and non-CHD. Sex specificity of the DMRs was tested by obtaining smoothed methylation values over DMRs from the Males Only comparison in female samples and over DMRs from the Females Only comparison in male samples, and PCA was performed as explained above.

Machine learning algorithms implemented through DMRichR were used to identify minimal DMRs for classifying samples as CHD or non-CHD (16). Random forest algorithms from the Boruta package (30) and support vector machine algorithms from the sigFeature package (31) were used to build binary classification models and rank the DMRs by importance for the feature selection analyses. Minimal DMRs were selected as those that were identified in both lists and were in the top 1%.

DS-CHD DMRs from Sex Combined, Females Only, and Males Only comparisons were overlapped by genomic coordinates using rtracklayer v1.54.0 (32) and GenomicRanges v1.46.1 (33), and the venn diagram was made with VennDiagram v1.7.3 (34) in R v4.1.3.

### 2.7 Enrichment testing and gene ontology from WGBS DMRs

DMRs from all comparisons were tested for enrichment in chromosome location compared to background regions using the Database for Annotation, Visualization and Integrated Discovery (DAVID), 2021 version (35,36). DMRs were tested for enrichment in genic (promoter, 5’UTR, exon, intron, 3’UTR, downstream, intergenic) and CpG (island, shore, shelf, open sea) contexts compared to background regions using DMRichR (16). The significance of genic and CpG annotations were tested using Fisher’s exact test and FDR correction. DMRs were mapped to genes on the hg38 genome using TxDb. Gene ontology enrichment was performed using rGREAT (37), with genomic coordinates of DMRs tested relative to background regions using the “oneClosest” rule.

### 2.8 Replication of WGBS CHD DMRs in independent DS newborn study

DMRs were tested for replication in a previously published DS NDBS WGBS dataset with 10 non-CHD (2 female, 8 male) and 11 CHD (6 female, 5 male) individuals (16). Unadjusted smoothed methylation values were calculated in replication dataset samples over DMR genomic coordinates from Sex Combined, Females Only, and Males Only comparisons using the getMeth function of the bsseq R package (27). Unpaired t-tests were calculated using the smoothed methylation values for replication CHD vs non-CHD samples and *p*-values were corrected by FDR using GraphPad Prism v9.4.1.

### 2.9 Comparison of WGBS CHD DMRs and background regions with DS vs TD NDBS samples

DS-CHD DMRs and background regions were tested for overlap with DMRs associated with DS in a previous epigenome-wide association study that included 21 DS (8 female, 13 male) and 32 TD (16 female, 16 male) NDBS samples with WGBS data (16). Unadjusted smoothed methylation values were calculated in replication dataset samples over DMR and background region genomic coordinates from Sex Combined, Females Only, and Males Only analyses using the getMeth function of the bsseq R package (27). Unpaired t-tests were calculated for DS vs TD using the smoothed methylation values of the replication dataset and *p*-values were corrected by the FDR method using GraphPad Prism v9.4.1. Potential differences between the proportions of DS-CHD DMRs and background regions that were significantly differentially methylated in DS vs TD or methylated in the same direction in DS vs TD as DS-CHD vs DS non-CHD were calculated using the z-test for two population proportions.

## 3. Results

### 3.1 Sample traits were not different in DS-CHD cases compared to DS non-CHD controls

We quantitated DNA methylation by EPIC array and WGBS in DNA isolated from NDBS from 86 individuals with DS, 45 of whom also had a CHD. Overall, our cohort had more females (*n* = 54, CHD = 27, non-CHD = 27) than males (*n* = 32, CHD = 18, non-CHD = 14) and a high proportion of Hispanic participants (*n* = 57 (63%)), compared to non-Hispanic white (*n* = 17 (19.8%)), non-Hispanic Asian (*n* = 8 (9.3%)), and non-Hispanic Black (*n* = 4 (4.7%)) participants; however, there were no significant differences for sex or race/ethnicity between DS-CHD and DS non-CHD newborns **(Table 1) (Supplemental Table S3)**. In addition, birthweight, gestational age, and age at blood collection did not differ significantly between DS-CHD and DS non-CHD newborns **(Table 1) (Supplemental Table S3)**.

**Table 1.**
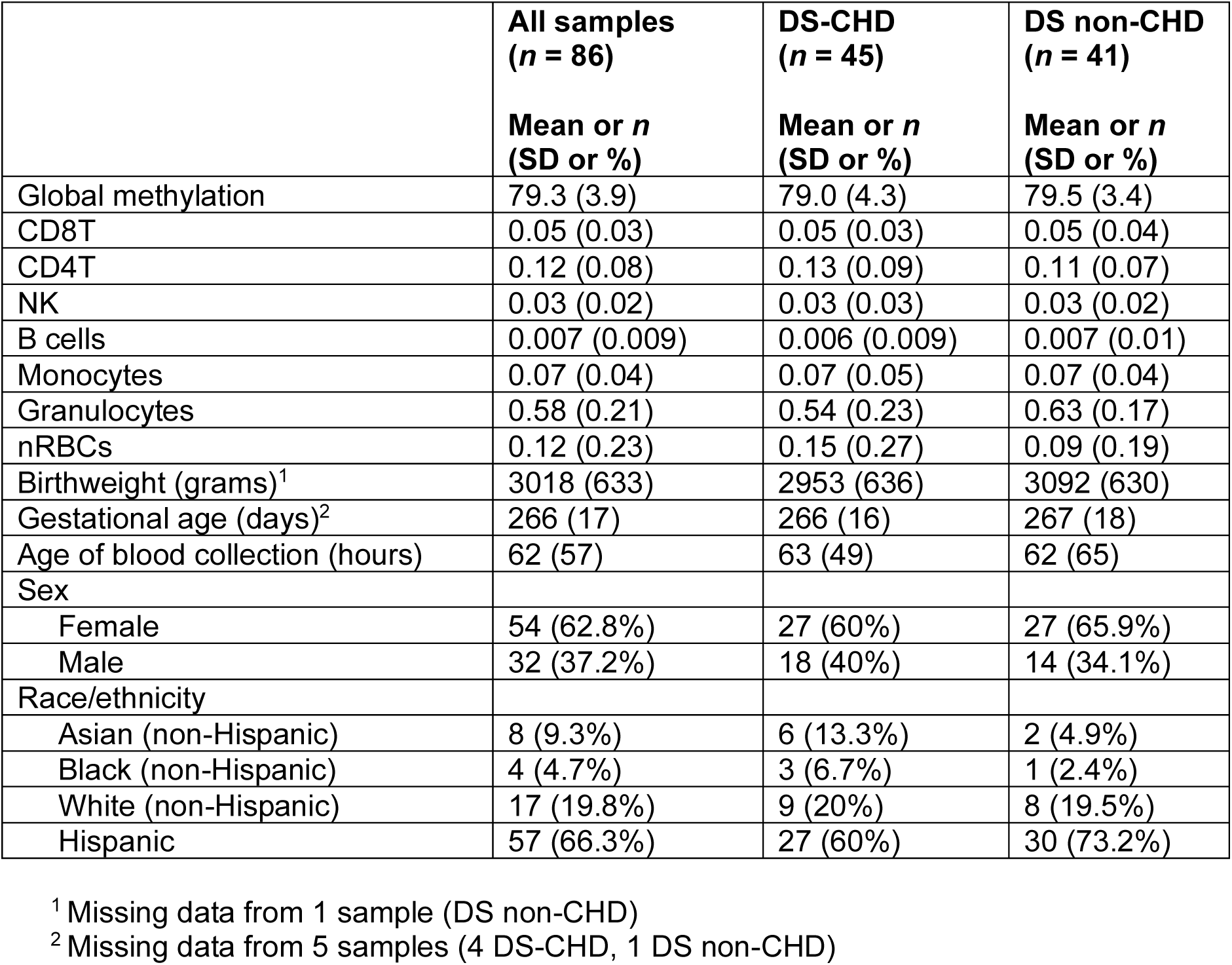
Sample traits in DS-CHD cases and DS non-CHD controls.

|  | **All samples**<br>**(*n* = 86)** | **DS-CHD**<br>**(*n* = 45)** | **DS non-CHD**<br>**(*n* = 41)** |
| --- | --- | --- | --- |
|  | **Mean or *n***<br>**(SD or %)** | **Mean or *n***<br>**(SD or %)** | **Mean or *n***<br>**(SD or %)** |
| Global methylation | 79.3 (3.9) | 79.0 (4.3) | 79.5 (3.4) |
| CD8T | 0.05 (0.03) | 0.05 (0.03) | 0.05 (0.04) |
| CD4T | 0.12 (0.08) | 0.13 (0.09) | 0.11 (0.07) |
| NK | 0.03 (0.02) | 0.03 (0.03) | 0.03 (0.02) |
| B cells | 0.007 (0.009) | 0.006 (0.009) | 0.007 (0.01) |
| Monocytes | 0.07 (0.04) | 0.07 (0.05) | 0.07 (0.04) |
| Granulocytes | 0.58 (0.21) | 0.54 (0.23) | 0.63 (0.17) |
| nRBCs | 0.12 (0.23) | 0.15 (0.27) | 0.09 (0.19) |
| Birthweight (grams)<sup>1</sup> | 3018 (633) | 2953 (636) | 3092 (630) |
| Gestational age (days)<sup>2</sup> | 266 (17) | 266 (16) | 267 (18) |
| Age of blood collection (hours) | 62 (57) | 63 (49) | 62 (65) |
| Sex |  |  |  |
| Female | 54 (62.8%) | 27 (60%) | 27 (65.9%) |
| Male | 32 (37.2%) | 18 (40%) | 14 (34.1%) |
| Race/ethnicity |  |  |  |
| Asian (non-Hispanic) | 8 (9.3%) | 6 (13.3%) | 2 (4.9%) |
| Black (non-Hispanic) | 4 (4.7%) | 3 (6.7%) | 1 (2.4%) |
| White (non-Hispanic) | 17 (19.8%) | 9 (20%) | 8 (19.5%) |
| Hispanic | 57 (66.3%) | 27 (60%) | 30 (73.2%) |

<sup>1</sup> Missing data from 1 sample (DS non-CHD)
<sup>2</sup> Missing data from 5 samples (4 DS-CHD, 1 DS non-CHD)

The estimated cell type proportions (CD8T, CD4T, NK, B cell, monocytes, granulocytes, nRBC) in newborn blood were highly variable across samples, particularly for nRBC and granulocyte proportions **(Supplemental Table S2)**. All cell types were positively correlated with one another, except nRBCs which were negatively correlated with all other cell types **(Supplemental Table S4) (Supplemental Figure S1)**. Most cell types (CD8T, CD4T, NK, monocytes, nRBC) were also significantly (unadjusted *p* <0.05) correlated with age of blood collection and all cell types were significantly correlated with WGBS global methylation levels, supporting the adjustment for cell types in our DMR analyses. Cell type proportions did not differ significantly between DS-CHD and DS non-CHD newborns overall, or in sex-stratified comparisons **(Table 1) (Supplemental Table S3)**.

### 3.2 WGBS of newborn blood DNA detects global hypomethylation in DS-CHD males compared to DS non-CHD males due to elevated nRBC proportions

To assess the reproducibility of EPIC array and WGBS methylation quantitation, we examined global CpG methylation levels from the two platforms and found that EPIC array beta values **(Supplemental Table S5)** were lower than WGBS global methylation values across all samples, but very strongly correlated (r = 0.9716, *p* <0.0001) **(Supplemental Figure S2)**. While a few samples had notably low global CpG methylation levels (<70% from WGBS), these samples were not removed from analysis because their other QC metrics were acceptable and their corresponding array beta values were also low, suggesting it was not a technical error.

Using WGBS data, we first assessed whether global CpG methylation levels differed between DS-CHD and DS non-CHD newborns. There was no significant difference overall, but when stratified by sex we found significant hypomethylation in DS-CHD males compared with DS non-CHD males (unadjusted *p* <0.05), a pattern that was not seen in females **(Figure 1A-B) (Supplemental Figure S3) (Supplemental Table S3)**. We confirmed by logistic regression that global methylation was the most predictive variable of CHD in males (*p* = 0.101), while CD4T cell proportion was the most predictive variable in females (*p* = 0.0681) **(Supplemental Table S6)**. Because nRBCs are known to have lower methylation levels than other cell types in blood (38) and their proportion in blood samples varies widely across individuals (39) in negative association with global methylation (38,40), we investigated the relationship between nRBC proportion and global methylation in our samples. In both females and males, nRBC proportion was significantly negatively correlated with global methylation levels **(Figure 1C-D) (Supplemental Table S4) (Supplemental Figure S1)** and was the most predictive variable of global methylation in linear regression models (females *p* <-2E16, males *p* =2.12E-9) **(Supplemental Table S6)**. Presence of CHD predicted global methylation levels in males (*p* =0.0618) much better than in females (*p* = 0.981), but addition of nRBC proportion as an adjustment covariate decreased the strength of this relationship (males, *p* =0.237) **(Supplemental Table S6)**.

**Figure 1.**
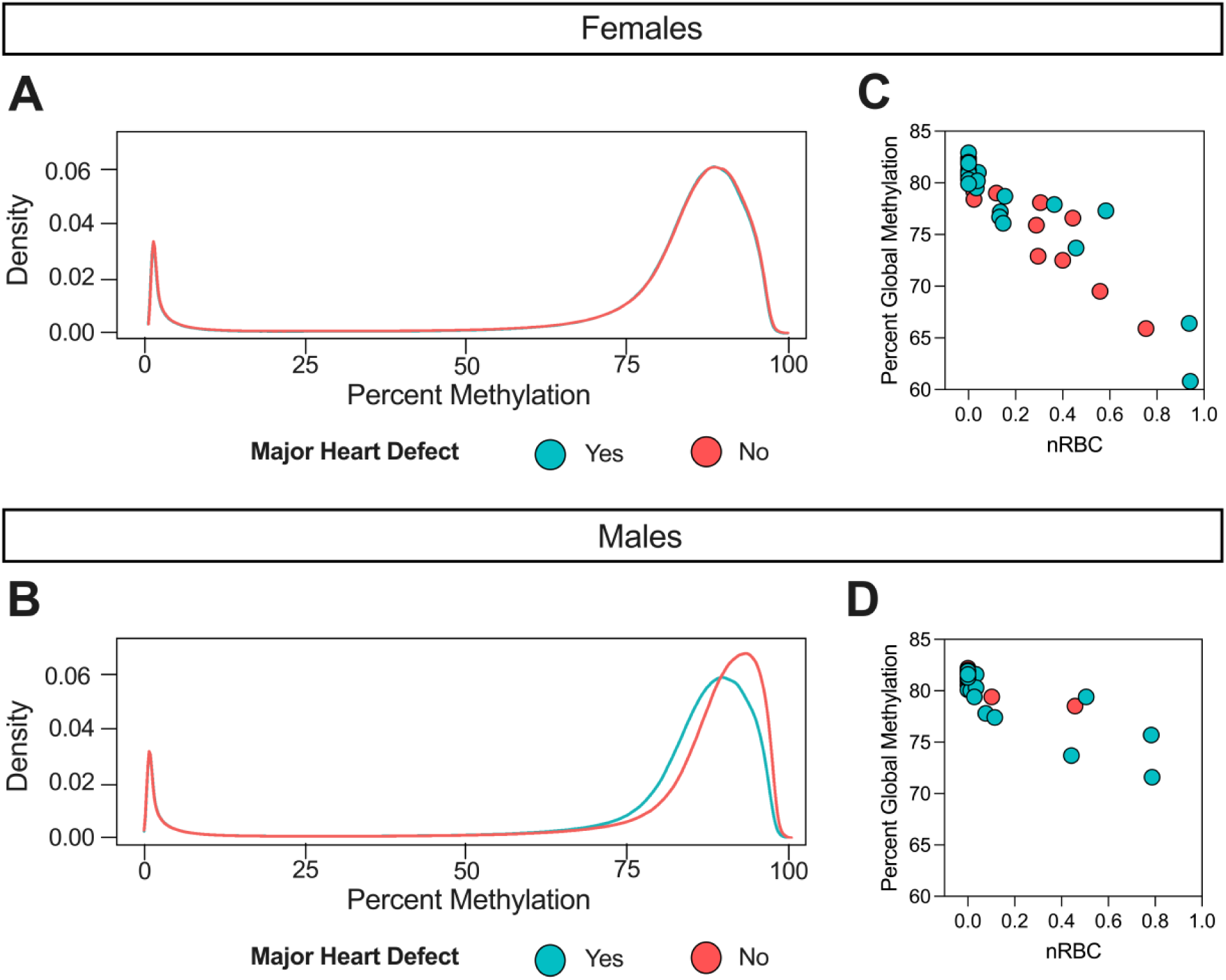
Global hypomethylation in DS-CHD males vs DS non-CHD males is driven by samples with high nRBC proportions. Density plot of average percent smoothed methylation in DS-CHD (Yes: blue) and DS non-CHD (No: red) in **A)** females (note that red and blue lines are overlapping) and **B)** males. Percent global methylation correlated with nRBC proportion in **C)** females (Pearson’s r = −0.93, *p* = 2.16E-24) and **D)** males (Pearson’s r = −0.84, *p* = 2.13E-9)

While proportion of nRBCs in the nucleated cell population is typically very low, with a median of 0 for the estimated nRBC proportions across samples in our study, we identified 12 out of 32 male samples with nRBC proportions >1%, of which 10 (83%) had a CHD **(Supplemental Table S2) (Supplemental Figure S3)**. In contrast, we identified 24/54 females with nRBC proportions >1% of which only 50% (n=12) had a CHD. In a sensitivity analysis, we removed five male samples that had notably high nRBC levels (>20%) and corresponding low global methylation levels and saw that global methylation in DS-CHD vs DS non-CHD male samples were no longer significantly different (*p* =0.1865) **(Supplemental Table S3)**. Females showed high interindividual variation in global methylation levels and nRBC proportions in both CHD and non-CHD groups, while CHD males showed much more variation (similar to females) than non-CHD males **(Supplemental Figure S3)**.

### 3.3 Sex-stratified DMRs distinguish DS-CHD from DS non-CHD samples better than sex-combined DMRs

Next, using WGBS data we investigated whether there were DMRs associated with DS- CHDs in Sex Combined, Females Only, and Males Only comparison groups to characterize both sex-specific and sex-independent patterns in DS-CHD methylation. We adjusted for confounding variables that were associated with global methylation (|r| >0.2): age of blood collection and all cell type proportions, as well as sex (specific for the Sex Combined comparison) **(Supplemental Figure S2) (Supplemental Table S4)**.

The Sex Combined comparison yielded 58 significant by permutation (*p* <0.05) DMRs **(Supplemental Table S7)**. In Females Only, we found 341 DMRs **(Supplemental Table S8)**, whereas in Males Only we found 3,938 DMRs **(Figure 2A) (Supplemental Table S9)**. Samples with low methylation levels across Males Only DMRs corresponded with those with low global methylation. In a sensitivity analysis excluding the five male samples with nRBC proportions >20%, we identified 2,474 Males Only DMRs **(Supplemental Table S10)**.

**Figure 2.**
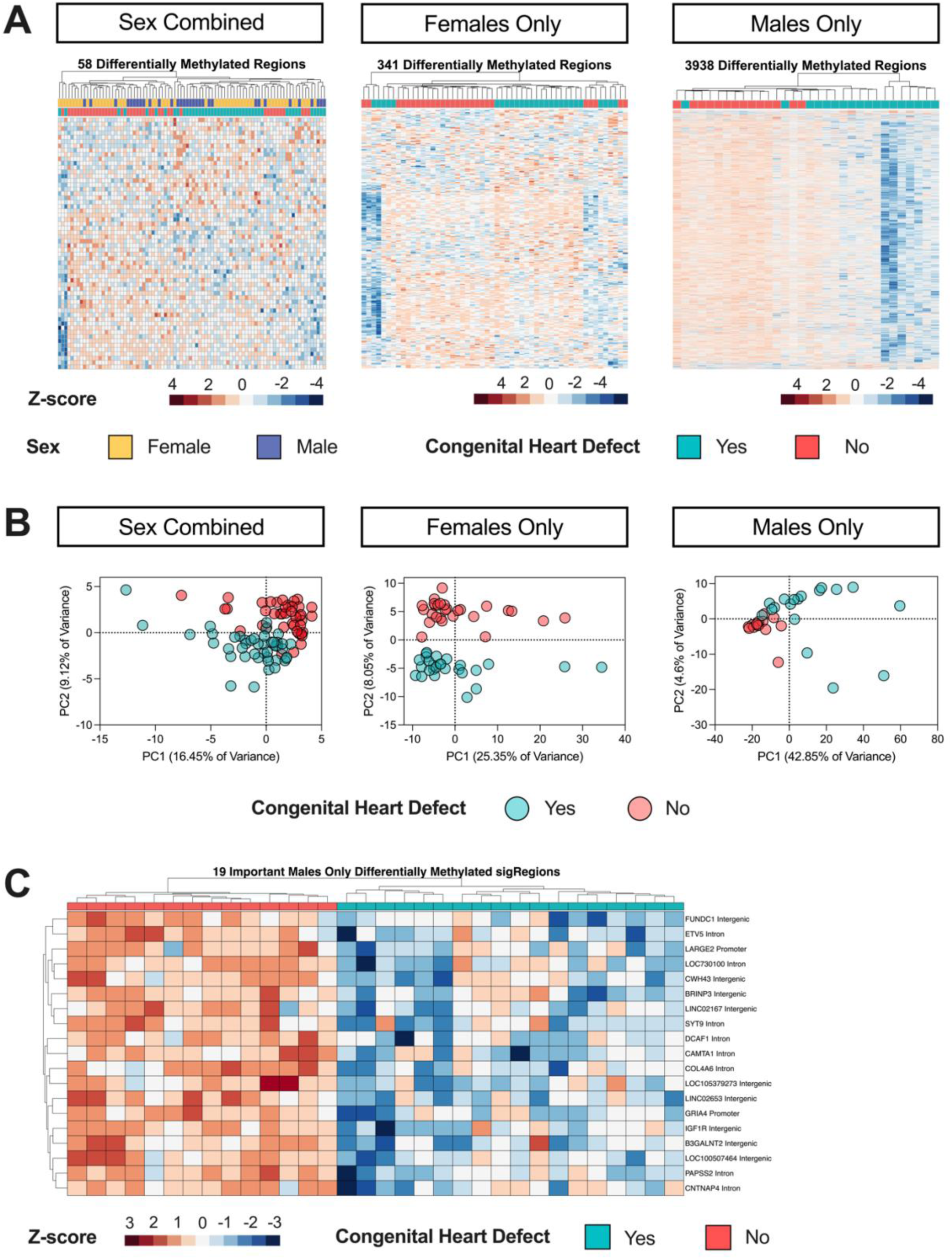
DMR profiles of CHD vs non-CHD in Sex Combined, Females Only, and Males Only comparisons within DS. **A)** Heatmaps of nominally significant (*p* <0.05) DMRs from DS-CHD vs DS non-CHD samples in Sex Combined, Females Only, and Males Only comparisons. All heatmaps show hierarchical clustering of Z-scores, which are the number of standard deviations from the mean of non-adjusted percent smoothed individual methylation values for each DMR. **B)** PCA analysis using the smoothed methylation values of all DMRs from the Sex Combined and Females Only comparisons and the 1000 most significant DMRs in the Males Only comparison. **C)** Hierarchical clustering heatmap of the machine learning feature selection analysis of the consensus DMRs from the Males Only comparison.

DMR hierarchal clustering and principal component analysis **(PCA)** showed that CHD and non-CHD samples did not separate completely, although sex stratification improved the distinction **(Figure 2A-B)**. Using machine learning feature selection, we identified a minimal set of 19 Males Only DMRs that could distinguish CHD from non-CHD samples **(Figure 2C) (Supplemental Table S11)**. The five male samples with high nRBCs and low methylation across DMRs did not have outlier methylation values across the 19 minimal DMRs, showing that the most predictive DMRs were not driven by outliers. In the Males

Only sensitivity analysis (with 5 samples with nRBC >20% removed), 13 minimal DMRs distinguished CHD from non-CHD, with four overlapping with those from the Males Only minimal selection using all samples: *DCAF1, LARGE2, LOC105379273, SYT9* **(Supplemental Table S11).** In the other comparisons, 6 Sex Combined and 3 Females Only DMRs were identified by the feature selection but could not cleanly distinguish CHD from non-CHD samples **(Supplemental Figure S4) (Supplemental Table S11)**.

In the replication dataset of WGBS from NDBS of 21 children with DS, 11 with CHD (6 females, 5 males) and 10 without CHD (2 females, 8 males) (16), we found 26 (46.4%) of the 56 Sex Combined DMRs that were covered in the replication study were methylated in the same direction in both groups **(Supplemental Table S12)**, while 161/329 (48.9%) of Females Only **(Supplemental Table S13)** and 2,229/3,938 (56.7%) of Males Only DMRs **(Supplemental Table S14)** were methylated in the same direction. Few DMRs were significantly differentially methylated (unadjusted *p* <0.05) in the replication dataset, with 2 Sex Combined, 9 Females Only, and 68 Males Only meeting this cutoff.

In DMR analysis using EPIC array data, there were no significant DMRs associated with DS-CHDs, likely due to the EPIC array only covering ∼3% of CpGs covered by WGBS (data not shown).

### 3.4 DS-CHD DMRs are sex-specific, with a small fraction overlapping across sexes

We next examined similarities and differences in DMRs across females and males. In the Sex Combined comparison, 60% of DMRs were hypomethylated in CHD compared to non-CHD samples, while 40% of Females Only DMRs and 96% of Males Only DMRs were hypomethylated **(Figure 3A)**. In our Males Only sensitivity analysis that removed samples with nRBC proportions >20%, 82% of DMRs were hypomethylated. To test the sex specificity of Females Only and Males Only DMRs, we analyzed the smoothed methylation values over DMRs from the Males Only comparison in female samples and from the Females Only comparison in male samples and found that CHD and non-CHD samples did not separate by PCA **(Supplemental Figure S5)**. Females and males also did not separate by hierarchal clustering or PCA in the Sex Combined comparison **(Figure 2A) (Supplemental Figure S6)**.

**Figure 3.**
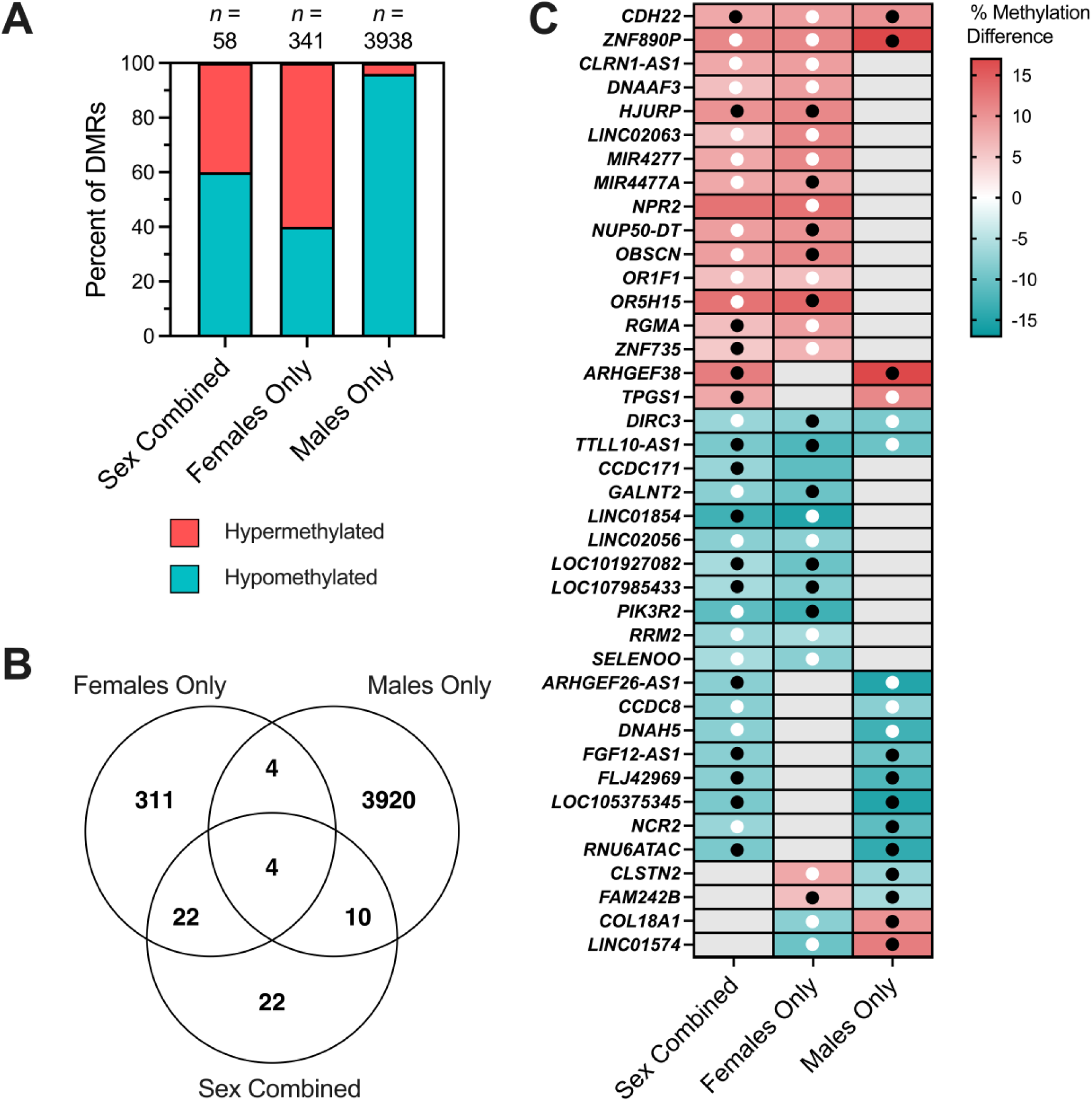
Overlapping CHD DMRs across Sex Combined, Female Only, and Male Only comparisons within DS. **A)** The percent of DMRs which were hypermethylated versus hypomethylated in each of the three comparisons. **B)** Venn diagram reflecting the numbers of unique and overlapping DMR genomic coordinates across the three comparisons. **C)** DS-CHD DMRs which overlap in two or more comparisons mapped to genes. Red indicates hypermethylation in CHD compared to non-CHD while blue represents hypomethylation, with stronger shades representing a greater percent methylation difference. Grey is used when a DMR was not called for that comparison. Black dots indicate methylation in the same direction in the discovery and replication datasets (10 non-CHD (2 female, 8 male) and 11 CHD (6 female, 5 male)) while white dots indicate methylation in the opposite direction in the two datasets. No dot means that the DMR genomic coordinates were not covered in the replication dataset.

DMR genomic coordinates from all comparisons were then overlapped to identify sex-specific vs sex-independent regions. Only 4 DMRs overlapped across all three comparisons, 22 across Sex Combined and Females Only comparisons, 10 across Sex Combined and Males Only comparisons, and 4 across Males Only and Females Only comparisons **(Figure 3B)**. All overlapping DMRs between comparison groups were methylated in the same direction except for the 4 overlapping between Females Only and Males Only comparisons (but not the Sex Combined comparison), which showed methylation in opposite directions **(Figure 3C)**. The 4 DMRs identified in all three comparisons mapped to *CDH22, ZNF890P, DIRC3*, and *TTLL10-AS1* genes.

### 3.5 DS-CHD DMRs are enriched for gene exons, CpG islands, and bivalent chromatin

CHD DMRs from Sex Combined, Females Only, and Males Only comparisons were analyzed for enrichment compared to background regions by distribution across chromosomes, genic and CpG contexts, histone marks, and chromatin states. In all three comparisons, DMRs were distributed throughout the genome **(Supplemental Figure S7)**, though Males Only DMRs showed significant enrichment (FDR <0.05) on chromosomes 2, 4, 5, 8, 18, and 21, while Females Only DMRs showed nominal enrichment (unadjusted *p* <0.05) on chromosomes 20, X, and 21 **(Supplemental Table S15)**. There was significant positive enrichment in all comparisons for gene exons and CpG islands **(Figure 4) (Supplemental Figures S8, S9) (Supplemental Table S16)**, as well as the transcriptionally repressive H3K27me3 histone mark and bivalent enhancers and transcription start sites based on chromatin states **(Supplemental Figures S10-S15)**. Sex differences were also observed, with significant positive enrichment for CpG shelves in the Females Only comparison and significant negative enrichment in the Males Only comparison. The Females Only DMRs also showed enrichment for H3K4me3, associated with active/poised chromatin, while the Males Only DMRs showed enrichment for H3K9me3, another repressive mark **(Supplemental Figure S13)**. Hypomethylated regions showed overall stronger enrichment for histone marks and chromatin states compared to hypermethylated regions **(Supplemental Figures S14-S15)**.

**Figure 4.**
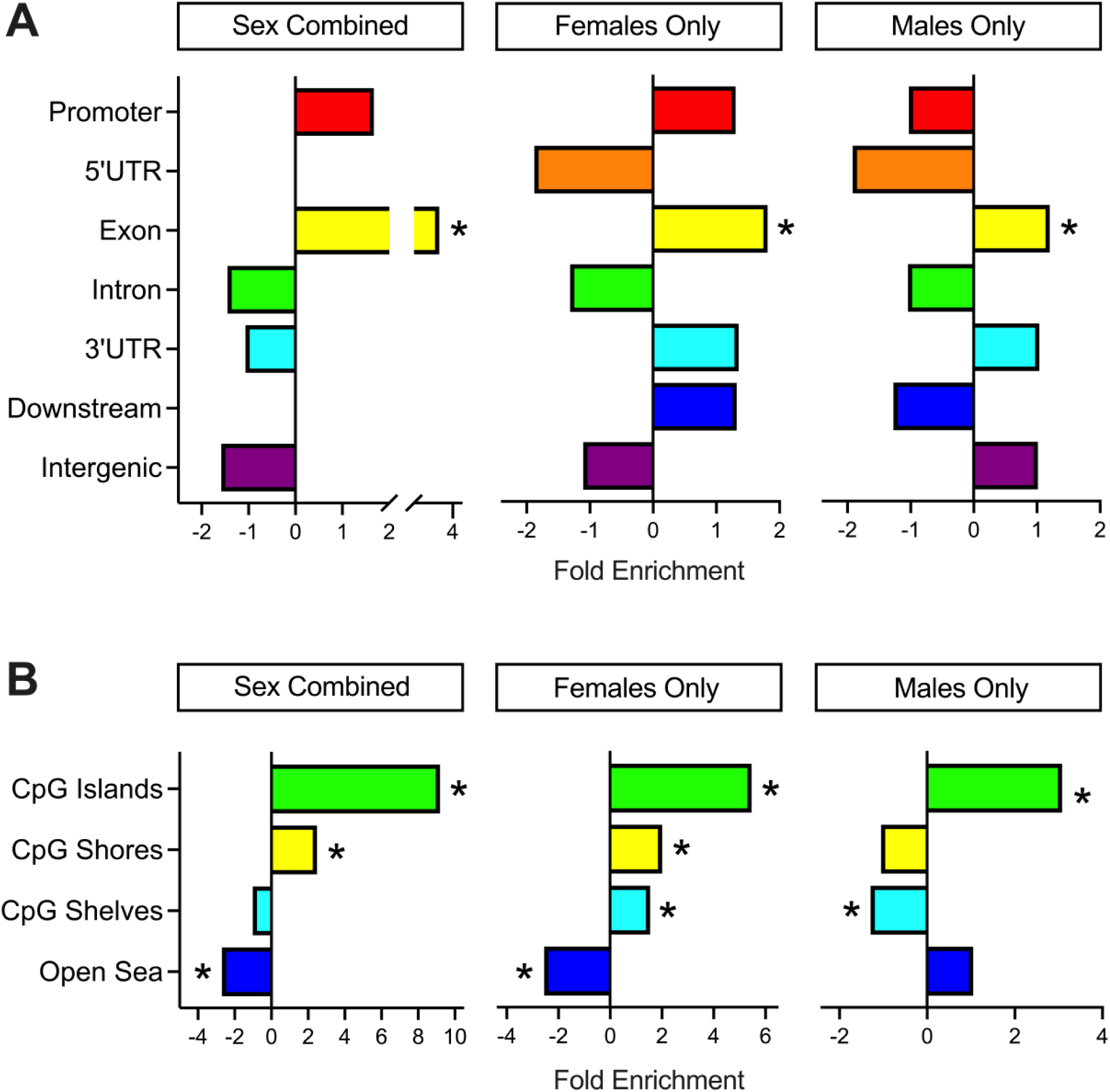
Annotation enrichments of CHD DMRs. **A)** Genic and **B)** CpG enrichments of all significant (*p* <0.05) DMRs from Sex Combined, Females Only, and Males Only comparisons. DMRs were compared to background regions for each comparison and significance was determined by the Fisher’s test and FDR correction. * = q < 0.05.

### 3.6 DS-CHD DMRs map to genes that are enriched for cardiac terms

DMRs mapped to genes were analyzed for enrichment across Gene Ontology terms (*p* <0.05) related to biological processes, cellular components, and molecular functions. All comparisons showed enrichment for heart-related terms, such as cardiac muscle contraction (Sex Combined) **(Figure 5)**, dorsal/ventral pattern formation, which includes formation of the embryonic heart tube (Females Only), and development of the septum primum, which divides the heart atrium into left and right and whose developmental failure can lead to AVSD (Males Only) **(Supplemental Figure S16) (Supplemental Tables S17- S19)**. Genes contributing to the heart-related terms included *FGF12, PIK3CA, TNNI3, PDE4D*, *ACVR1, GATA4,* and others **(Table 2)**. Enriched terms also included immune-related biological processes, such as platelet activation and innate immune response (Sex Combined) **(Figure 5)**.

**Figure 5.**
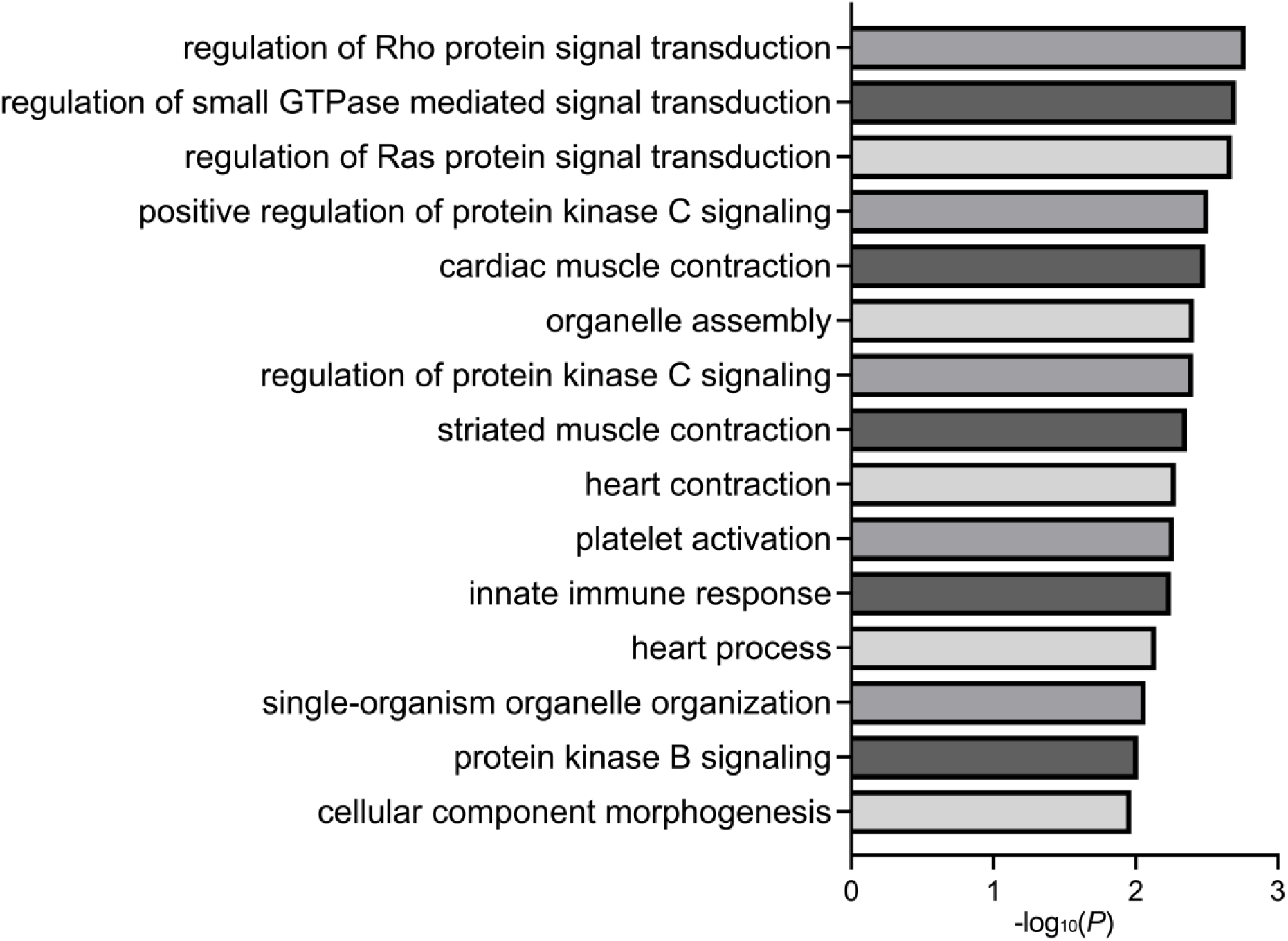
Gene ontology enrichments. Bar plot of the fifteen most significant GO enrichments for biological processes in DS-CHD versus DS non-CHD DMRs from the Sex Combined comparison.

**Table 2.**
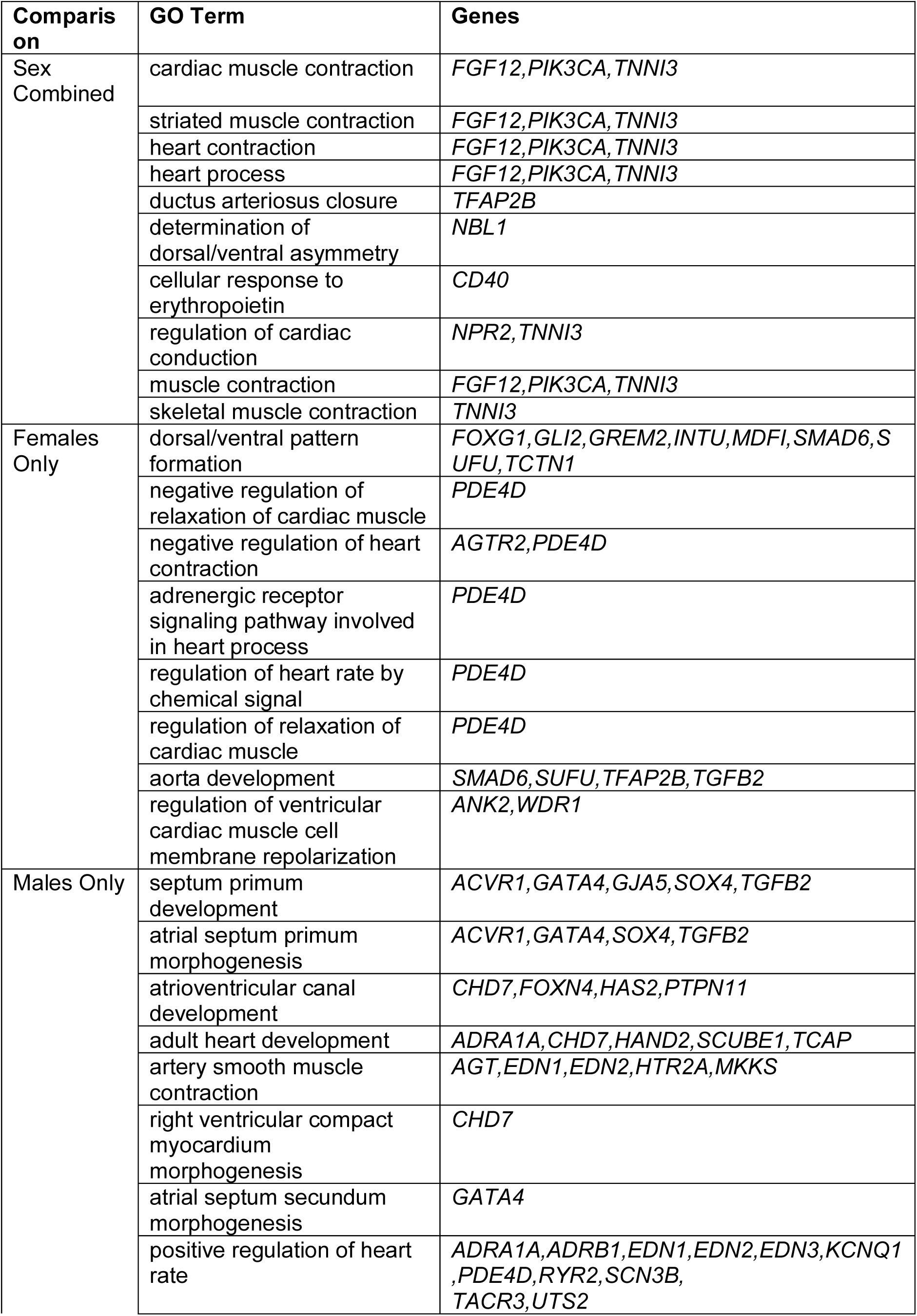
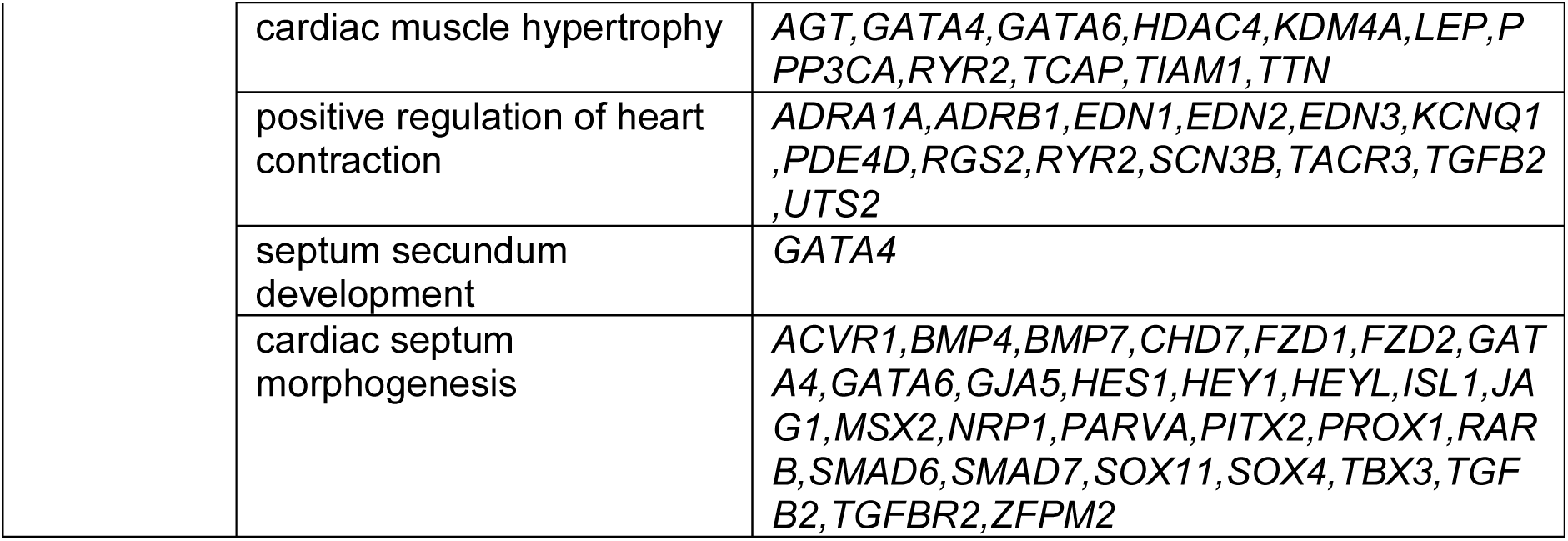
Heart-related biological processes identified from DMR Gene Ontology.

### 3.7 DS-CHD DMRs are also differentially methylated in DS vs typical development NDBS

DS-CHD DMRs were tested for comparison in previously published DS vs TD NDBS WGBS samples (16) to evaluate the hypothesis that if DS-CHD is a more severe form of DS, CHD DMRs should be partially shared with DS vs TD DMRs **(Table 3)**. Of the 58 Sex Combined CHD DMRs, 16 (27.6%) were significantly differentially methylated (*p* <0.05) in DS vs TD samples **(Supplemental Table S20)**, 9 of which (56.3%) were methylated in the same direction in DS vs TD samples as DS-CHD vs DS non-CHD samples. Of Females Only DMRs, 42/341 (12.3%) were significantly differentially methylated (*p* <0.05) in DS vs TD, with 28 (66.7%) methylated in the same direction **(Supplemental Table S21)** and of Males Only DMRs, 602/3,938 (15.3%) were significantly differentially methylated (*p* <0.05) in DS vs TD, with 528 (87.7%) methylated in the same direction **(Supplemental Table S22)**. These numbers decreased in a sensitivity analysis with the Males Only DMRs generated with five samples with nRBC >0.2 removed, where 334/2,454 (13.6%) covered DMRs were significantly (*p* <0.05) differentially methylated in DS vs TD male samples, of which 248/334 (74.3%) were methylated in the same direction. For all three comparisons, there was a trend towards more CHD DMRs being significantly differentially methylated in DS vs TD samples compared to background regions (z-test for two population proportions, Sex Combined *p* =0.08364, Females Only *p* =0.0536, Males Only *p* =0.0601) **(Figure 6A).** In Males Only, significantly more DMRs were methylated in the same direction in DS vs TD as DS-CHD vs DS non-CHD compared to background regions (z-test for two population proportions, *p* < 0.00001), though this was not true for Sex Combined or Females Only CHD DMRs **(Figure 6B)**. Of DMRs that were significantly differentially methylated (*q* <0.05) in DS vs TD samples, 5/9 (55.6%) Sex Combined, 6/8 (75%) Females Only, and 15/16 (93.85%) Males Only DMRs were hypomethylated in DS compared to TD samples. With the exception of an exon in *ZNF735*, which was significantly hypermethylated (*q* <0.05) in both the Sex Combined and Females Only DS vs TD comparisons, all DMRs were specific to one comparison **(Figure 6C)**.

**Figure 6.**
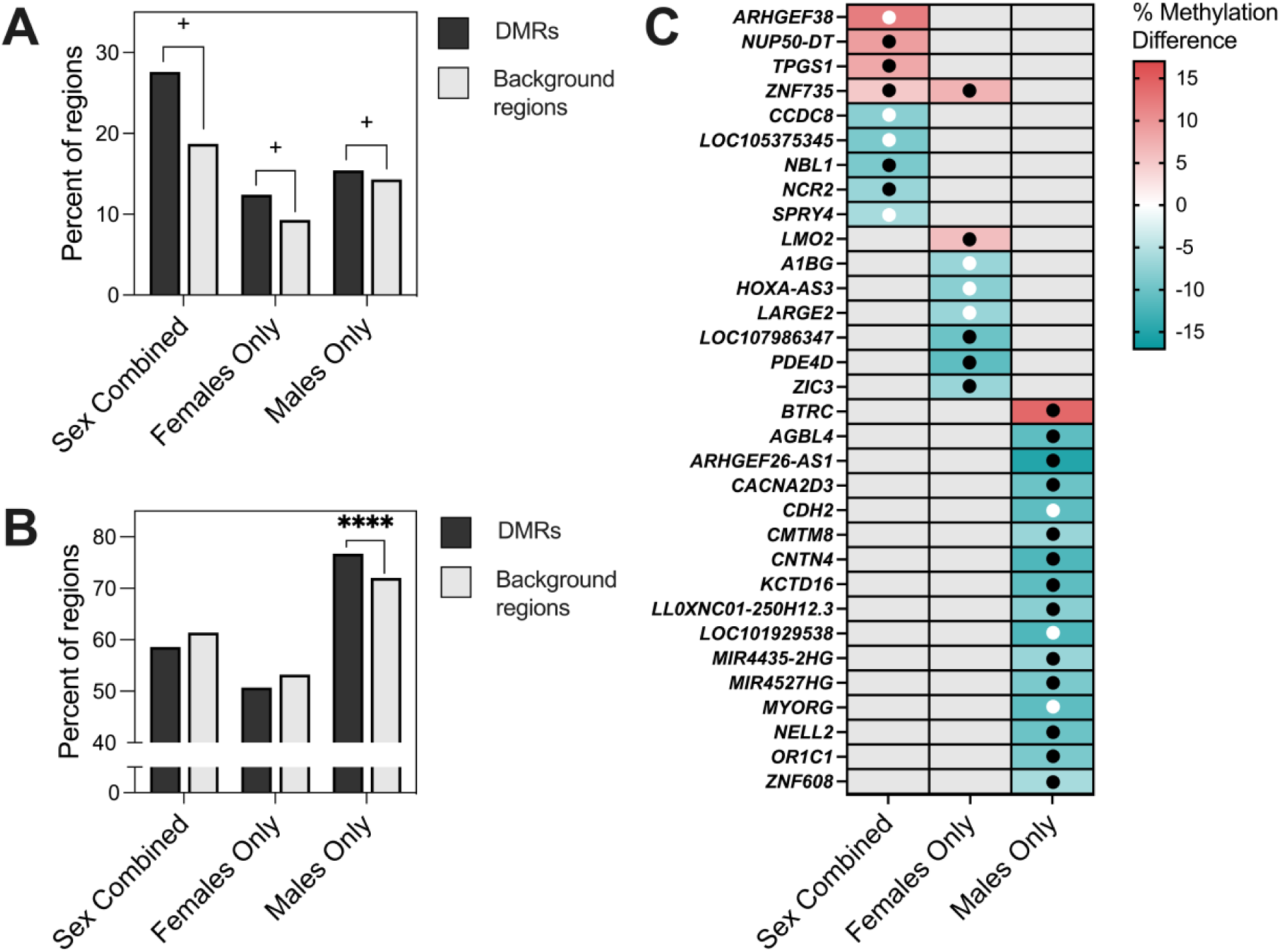
Comparison of DS-CHD DMRs with DS vs TD samples. **A)** Percent of DS-CHD DMRs and background regions that were significantly differentially methylated in DS vs TD samples. Z-test for two population proportions, Sex Combined (z =1.7343, two-tailed *p* = 0.08364) Females Only (z =1.93, two-tailed *p* =0.0536), Males Only (z =1.8808, two-tailed *p* =0.0601). + = *p* <0.1. **B)** Percent of DS-CHD DMRs that were methylated in same direction in DS vs TD as in DS-CHD vs DS non-CHD. Z-test for two populations proportions, Sex Combined (z =-0.4274, two-tailed *p* =0.6672), Females Only (z =-0.8936, two-tailed *p* =0.37346), Males Only (z = 6.5357, two-tailed *p* <0.00001). **** = *p* <0.00001. **C)** Heatmap showing DS-CHD DMRs that were significant (q < 0.05) in DS vs TD samples mapped to genes. Red indicates hypermethylation in CHD compared to non-CHD while blue represents hypomethylation, with stronger shades representing a greater percent methylation difference and gray meaning that that DMR was not significant for that comparison. Black dots indicate that methylation is in the same direction for DS vs TD as DS-CHD vs DS non-CHD while white dots indicate methylation is in the opposite direction.

**Table 3.** Significance and direction of CHD DMRs and background regions in DS vs TD samples.

|  | DMRs | Background Regions |
| --- | --- | --- |
| **Sex Combined** | *n* (%) | *n* (%) |
| Total | 58 | 5,363 |
| Omitted | 0 | 28 |
| $p < 0.05$ | 16 (27.6) | 995 (18.7) |
| $p \geq 0.05$ | 42 (72.4) | 4,340 (81.3) |
| Same direction | 34 (58.6) | 3,274 (61.4) |
| Opposite direction | 24 (41.4) | 2,061 (38.6) |
| **Females Only** | *n* (%) | *n* (%) |
| Total | 341 | 11,998 |
| Omitted | 2 | 147 |
| $p < 0.05$ | 42 (12.4) | 1,101 (9.3) |
| $p \geq 0.05$ | 297 (87.6) | 10,750 (90.7) |
| Same direction | 172 (50.7) | 6,304 (53.2) |
| Opposite direction | 167 (49.3) | 5,547 (46.8) |
| **Males Only** | *n* (%) | *n* (%) |
| Total | 3,938 | 127,819 |
| Omitted | 26 | 1,413 |
| $p < 0.05$ | 602 (15.4) | 18,099 (14.3) |
| $p \geq 0.05$ | 3,310 (84.6) | 108,307 (85.7) |
| Same direction | 3,001 (76.7) | 90,954 (72.0) |
| Opposite direction | 911 (23.3) | 35,452 (28.0) |

Same direction indicates methylation is in same direction (hypo or hyper) in DS vs TD as in DS-CHD vs non-CHD

## 4. Discussion

This is the largest study to date to investigate epigenetic variation associated with CHDs in individuals with DS. Although non-syndromic CHDs have been widely studied, there has been relatively little research into the etiology and biomarkers of CHDs in individuals with DS, despite nearly half of the DS population presenting this phenotype (2–5). To address this gap, we assessed DS-CHD methylation in DNA isolated from NDBSs, an understudied and accessible biospecimen that enables the analysis of epigenomic changes during the *in utero* and perinatal periods that are associated with phenotypic traits of interest. We confirmed the reproducibility of DNA extraction and bisulfite conversion from NDBSs by finding a high correlation (r = 0.9716) between global methylation levels from WGBS and EPIC array, which used different punches from the same blood spots.

Although newborn blood typically shows ∼79-84% global CpG methylation from WGBS (16,40), we found a range of 60.8-82.9% in our NDBS DNA. The samples with notably low WGBS global methylation also had low EPIC array beta values and passed other QC metrics, indicating that technical errors do not explain the result, although we cannot exclude non-biological causes. Previous studies have found a trend towards global hypomethylation in DS compared to TD NDBS (16), which may explain our findings. We further found that global methylation was lower in DS-CHD males compared to DS non-CHD males, though this relationship was not found in females. Global methylation was strongly negatively correlated with, and predicted by, nRBC proportion in both sexes. Although nRBCs typically constitute a very small proportion of nucleated cells, we found nRBC proportions ranged widely in both sexes and were evenly spread amongst CHD and non-CHD females but in males, 10/12 samples with nRBC >1% were CHD positive. This discrepancy in the relationship between high nRBCs and CHD in males versus females likely explains the association between CHD and global hypomethylation in males and the lack of such association among females, given the strong effects of nRBC proportions on global methylation levels in DS newborns (17). The etiology of the male-specific association between high nRBCs and CHD in newborns with DS remains to be determined.

Previous studies have reported high nRBC levels in DS newborns with pulmonary hypertension (41), as well as in hypoxic-related pregnancy situations, such as preeclampsia, maternal obesity and diabetes, maternal smoking, and prenatal exposure to infection (42–48). Increased nRBC counts are thought to follow fetal hypoxemia through elevated erythropoietin **(EPO)**, a hormone that stimulates production of erythrocytes (red blood cells) in an effort to increase oxygen delivery to tissues (49,50). Interestingly, EPO is higher in children with DS-CHD compared to non-syndromic CHD (51). Because CHDs reduce cerebral oxygen (52) and may induce fetal hypoxemia (53), high nRBC proportions may be more common in individuals with CHDs and, in particular, in DS newborns with CHDs given the placental abnormalities seen in fetuses with trisomy 21 (PMID: 31683073). However, we could not confirm this hypothesis with our sample size. Moreover, nRBC proportions were estimated from DNA methylation array data, rather than using actual cell counts, and cell type deconvolution in individuals with DS may be confounded by the presence of blast cells that are common in DS and not accounted for in the analysis (17). Cell composition deconvoluted from DNA methylation arrays has been previously reported to be altered in DS blood compared to non-DS blood (54). Further, we previously reported a positive relationship between high nRBC proportions in newborns with DS and presence of somatic *GATA1* mutations, indicative of transient abnormal myelopoiesis **(TAM)** or silent TAM, and it is possible that the relationship between CHD and global hypomethylation in males may be confounded by this preleukemic condition (17). Understanding of the complex relationships between CHDs in DS, global methylation, cell type proportions, sex, and fetal hypoxia would benefit from further investigation.

In our WGBS regional analysis, we found over 10-fold the number of CHD-associated DMRs in DS males than in females, and even fewer DMRs in the Sex Combined analysis. Reflecting our finding of global hypomethylation in CHD males, 96% of Males Only DMRs were hypomethylated, a pattern not seen in the Females Only or Sex Combined analyses. All DMRs were corrected for confounding factors including cell type proportions, suggesting that nRBC levels were not fully responsible for the notable proportion of hypomethylated DMRs in males, although we cannot rule out residual confounding due to nRBCs or unmeasured traits related to nRBCs. Additionally, removing the five male samples with nRBC proportions > 20% resulted in 82% hypomethylated DMRs, suggesting that these five samples alone were not driving the signature of hypomethylation in DS-CHD males. Some DMRs from all comparisons were also differentially methylated in DS vs TD samples, and in males, a significantly higher proportion of DMRs were methylated in the same direction in DS vs TD and DS-CHD vs DS non-CHD compared to background regions. These results suggest that male DS patients with CHD may represent a more severe epigenomic signature than is observed for DS versus TD, although this may also reflect the higher nRBC proportions that have been reported in newborns with DS than in TD newborns (Muskens 2021). In contrast, female DS cases with CHD are somewhat epigenetically distinct from female and male DS cases without CHD. Response to hypoxia may play a role in these differences. DS newborns, even those without CHDs, experience more hypoxemia events than newborns with TD (55), and CHDs further induce fetal hypoxemia (53). A wide variety of sex differences have been observed in response to hypoxia in both humans and animal models (56,57), including differences in gene expression profiles of female vs male mice in cardiac adaptive responses to hypoxia (58). These sex-specific responses to hypoxia may be reflected in the methylome, which is known to be influenced by gestational hypoxia (59) (reviewed in (60)). Identification of genes and pathways whose methylation and/or gene expression is altered in DS, CHDs, and hypoxia may help elucidate the sex specificity of molecular mechanisms related to DS-CHD.

Although we did not find any significant DMRs associated with DS-CHD after FDR-correction, the nominally significant DMRs were enriched for genes implicated in cardiac processes, suggesting that at least some of the DMRs may reflect true epigenetic mechanisms associated with DS-CHD development. In particular, Males Only DMRs selected by machine learning feature selection were able to distinguish CHD from non-CHD samples and frequently mapped to genes associated with CHDs or cardiomyopathies, including *FUNDC1* (61), *ETV5* (62,63), *SYT9* (64), *CAMTA1* (65), *GRIA4* (66), and *IGF1R* (67–70). Additionally, DMRs that contributed to enrichment for heart-related gene ontology terms included *TNNI3*, a cardiac-specific gene that codes for cardiac troponin I, whose absence leads to severe pediatric cardiomyopathy (71), and *GATA4,* which encodes a member of the GATA family of zinc finger transcription factors, is essential for mammalian cardiac development, and whose sequence variants have been identified in individuals with CHDs (72). Whether the differential methylation in the genes we identified plays an etiologic role or reflects epigenomic effects downstream of the development of CHDs remains to be determined.

While this is, to our knowledge, the largest DNA methylation study of CHDs in DS, our sample size of 86 DS newborns may still have limited our ability to detect genome-wide significant (*q* <0.05) DMRs. Additionally, only around half of DMRs in all comparisons were methylated in the same direction in the discovery and replication groups, potentially due to the very small sample sizes and absence of confounding variable data to use for correction in the replication group, as well as high interindividual variation in methylation. The genes to which our DMRs mapped did not heavily coincide with those identified in previous epigenetic studies of DS-CHD (73,74), likely because those studies included small numbers of DS subjects, used non-NDBS biospecimens assayed with array-based methods, which do not have good coverage over the regions we detected using WGBS, and did not account for cell type heterogeneity. One exception to this is that we identified a DMR in the Males Only comparison that mapped to *SHC3*, a gene that was differentially expressed in DS individuals with an endocardial cushion CHD (73). The DS field would benefit from further studies into the etiology and biomarkers of phenotypes common in the DS population, including CHDs.

## 5. Conclusions

Overall, this study presents the largest investigation of epigenetic variation associated with CHDs in individuals with DS. We identified sex-specific global and regional methylation differences in DS-CHD vs DS non-CHD newborns. Specifically, in males we found that newborns with DS-CHD were globally hypomethylated compared to DS newborns without CHD, a finding that appeared to be driven by differences in nRBC proportions between the two groups. At the regional level, the majority of CHD DMRs identified by sex stratification did not overlap by genomic coordinates, suggesting sex differences in the molecular signature of CHDs in DS. Gene ontology analysis of DMRs from both sexes revealed enrichment in pathways related to the heart, and some DS-CHD DMRs were also differentially methylated in DS vs TD samples. Our results provide insight into the development of CHDs in newborns with DS, pointing to sex-specific differences that warrant further investigation, and suggest that DNA methylation may serve as a useful biomarker for investigating the variability of clinical features within the genetic disorder of DS.

## Data Availability

This study used biospecimens from the California Biobank Program. Any uploading of genomic data (including genome‐wide DNA methylation data) and/or sharing of these biospecimens or individual data derived from these biospecimens has been determined to violate the statutory scheme of the California Health and Safety Code Sections 124980(j), 124991(b), (g), (h), and 103850 (a) and (d), which protect the confidential nature of biospecimens and individual data derived from biospecimens. Should we be contacted regarding individual-level data contributing to the findings reported in this study, inquiries will be directed to the California Department of Public Health Institutional Review Board to establish an approved protocol to utilize the data, which cannot otherwise be shared peer-to-peer.

## List of abbreviations

DS: Down syndrome
CHD: Congenital heart defect
AVSD: Atrioventricular septal defect
WGBS: Whole-genome bisulfite sequencing
NDBS: Newborn dried blood spot
TD: Typical development
DMR: Differentially methylated region
QC: Quality control
nRBC: Nucleated red blood cell
PCA: Principal component analysis
EPO: Erythropoietin
TAM: Transient abnormal myelopoiesis

## DECLARATIONS

### Ethics approval and consent to participate

This study was approved by Institutional Review Boards at the California Health and Human Services Agency, University of Southern California, and University of California Davis.

### Consent for publication

Deidentified NDBS were obtained from the California Biobank Program (SIS request number 572), with a waiver of consent from the Committee for the Protection of Human Subjects of the State of California.

### Availability of data and materials

This study used biospecimens from the California Biobank Program. Any uploading of genomic data (including genome-wide DNA methylation data) and/or sharing of these biospecimens or individual data derived from these biospecimens has been determined to violate the statutory scheme of the California Health and Safety Code Sections 124980(j), 124991(b), (g), (h), and 103850 (a) and (d), which protect the confidential nature of biospecimens and individual data derived from biospecimens. Should we be contacted regarding individual-level data contributing to the findings reported in this study, inquiries will be directed to the California Department of Public Health Institutional Review Board to establish an approved protocol to utilize the data, which cannot otherwise be shared peer- to-peer.

Code is available at https://github.com/juliamouat/DownSyndrome_CongenitalHeartDefect_DNAmethylation

### Authors’ contributions

BIL, AJD, and JML designed the study. AJD and JML supervised the project. AJD, PJL, JPW, and JMS prepared data. SSM performed DNA extractions. SL performed array analysis. JSM performed bioinformatic analyses. JSM, AJD, and JML interpreted results. JSM drafted the manuscript and made figures and tables. PJL, AJD, and JML revised the manuscript. All authors reviewed and approved the final manuscript.

## Acknowledgements

A subset of biospecimens and/or data used in this study were obtained from the California Biobank Program at the California Department of Public Health (CDPH), SIS request number 572, in accordance with Section 6555 (b), 17 CCR. The CDPH is not responsible for the results or conclusions drawn by the authors of this publication.

We thank Robin Cooley and Steve Graham (Genetic Disease Screening Program, CDPH) for their assistance and expertise in the procurement and management of NDBS specimens. We thank the DNA Technologies and Expression Analysis Core at the UC Davis Genome Center for library preparation and WGBS.

