## Supplemental Figures for "Epigenomic signature of major congenital heart defects in newborns with Down syndrome"

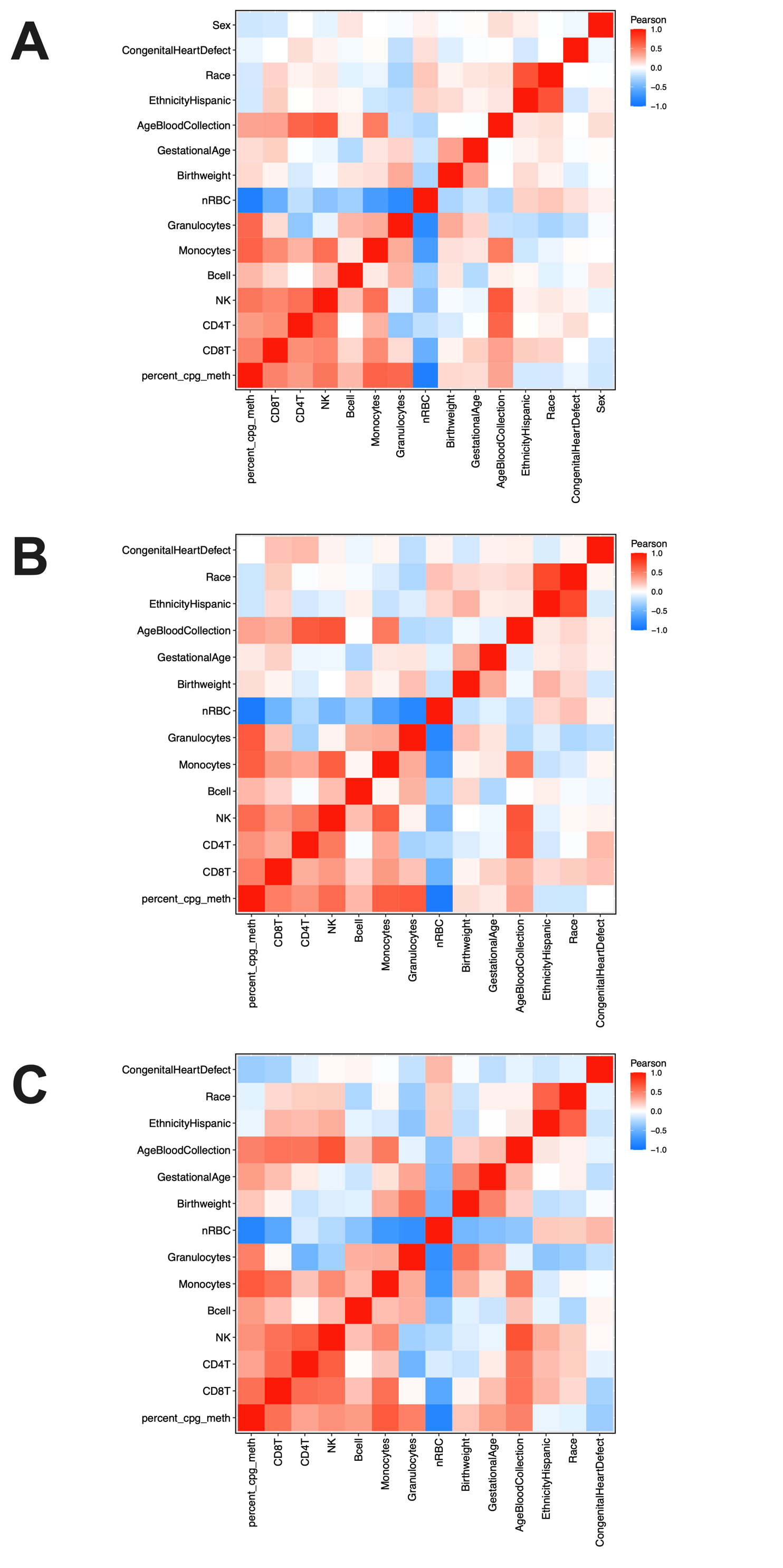

**Supplemental Figure S1.** Sample trait correlations in **A)** all samples, **B)** females only, and **C)** males only by Pearson correlation

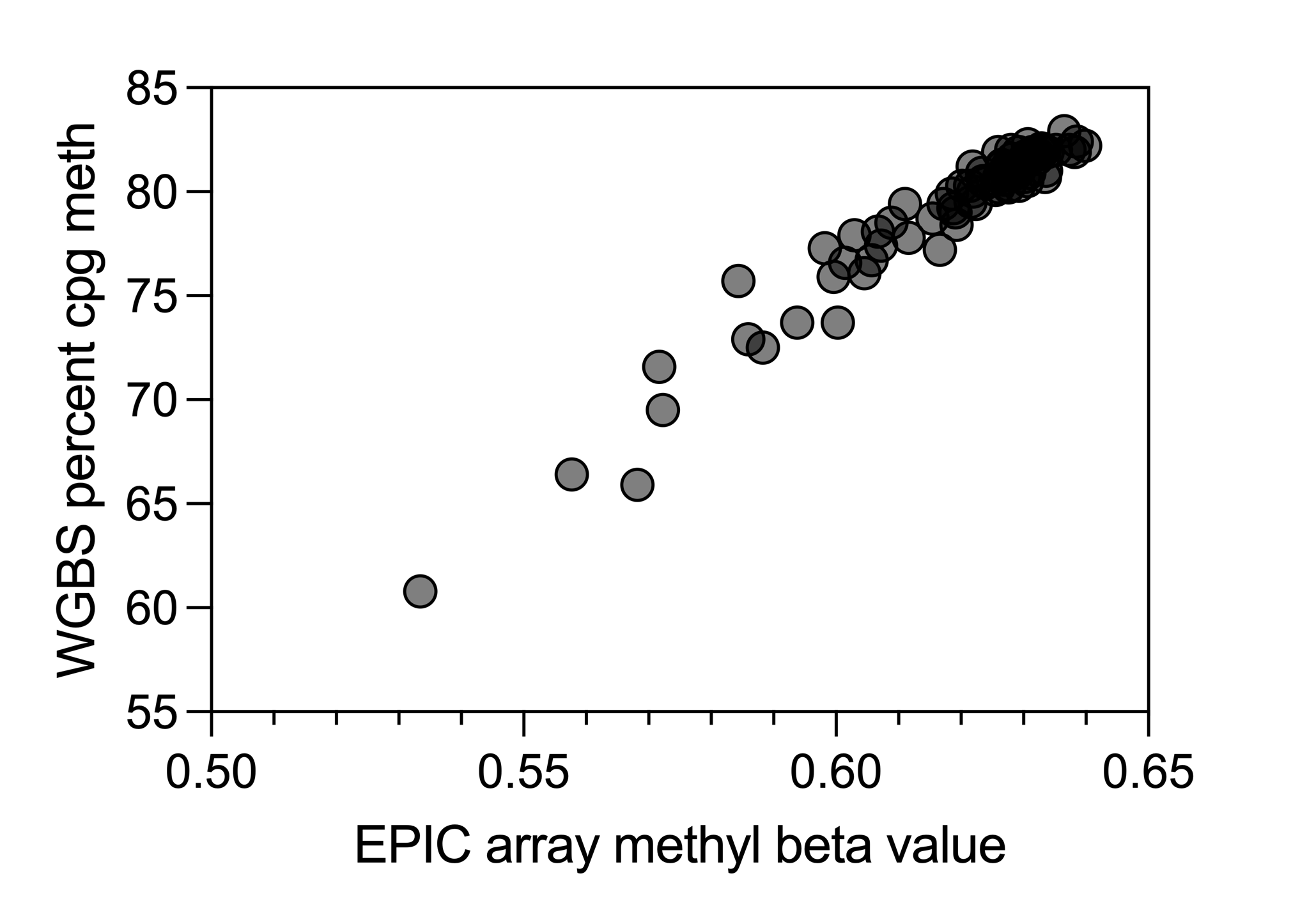

**Supplemental Figure S2**. Global methylation values from WGBS are highly correlated with those from the EPIC array. Pearson’s r =0.9716, *p* <0.0001

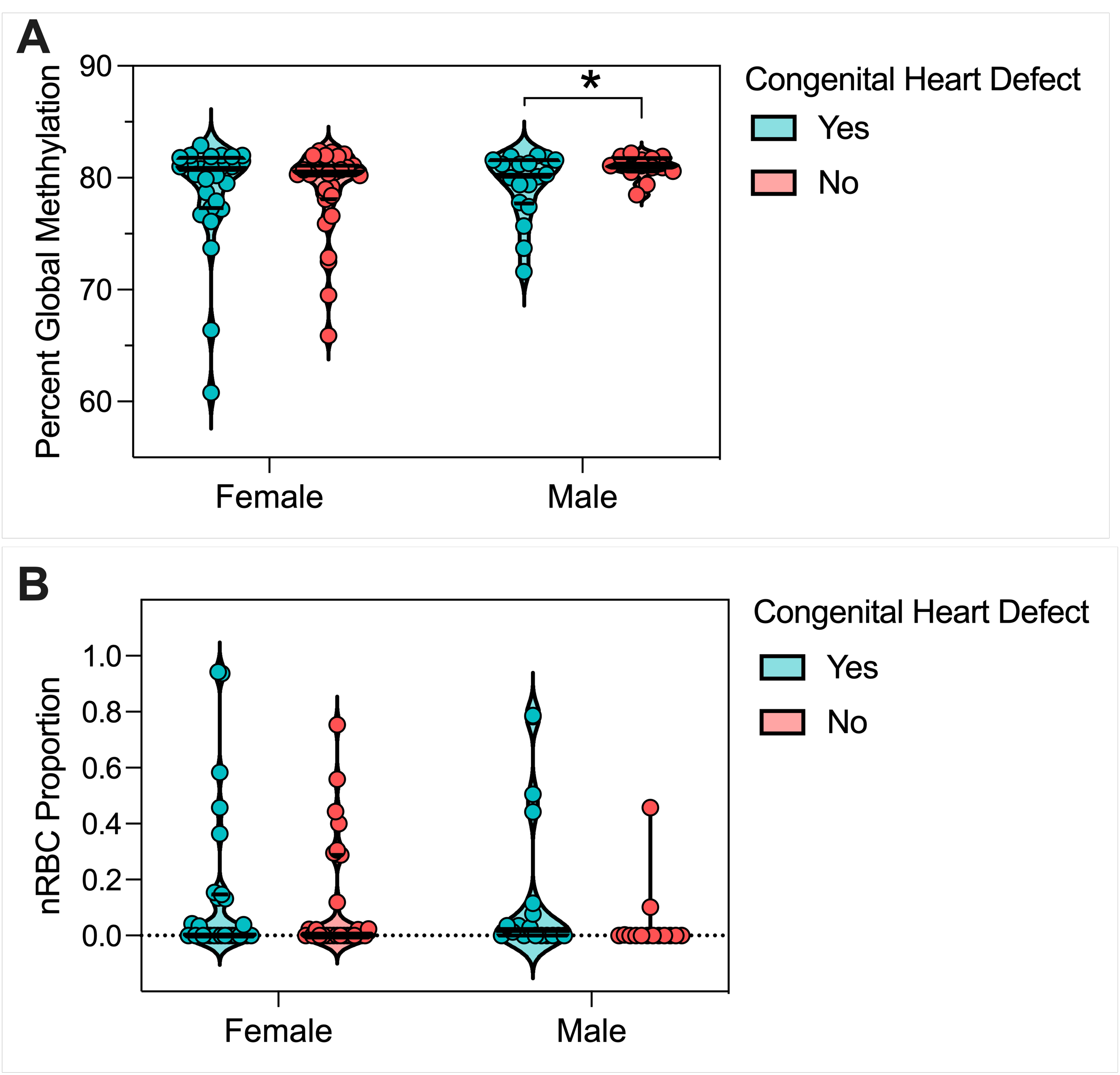

**Supplemental Figure S3.** Violin plots with all points shown for **A)** global methylation and **B)** nRBC proportion in females and males with DS CHD (Yes) and DS non-CHD (No). * = *p* <0.05 in Welch’s unequal variances *t*-test

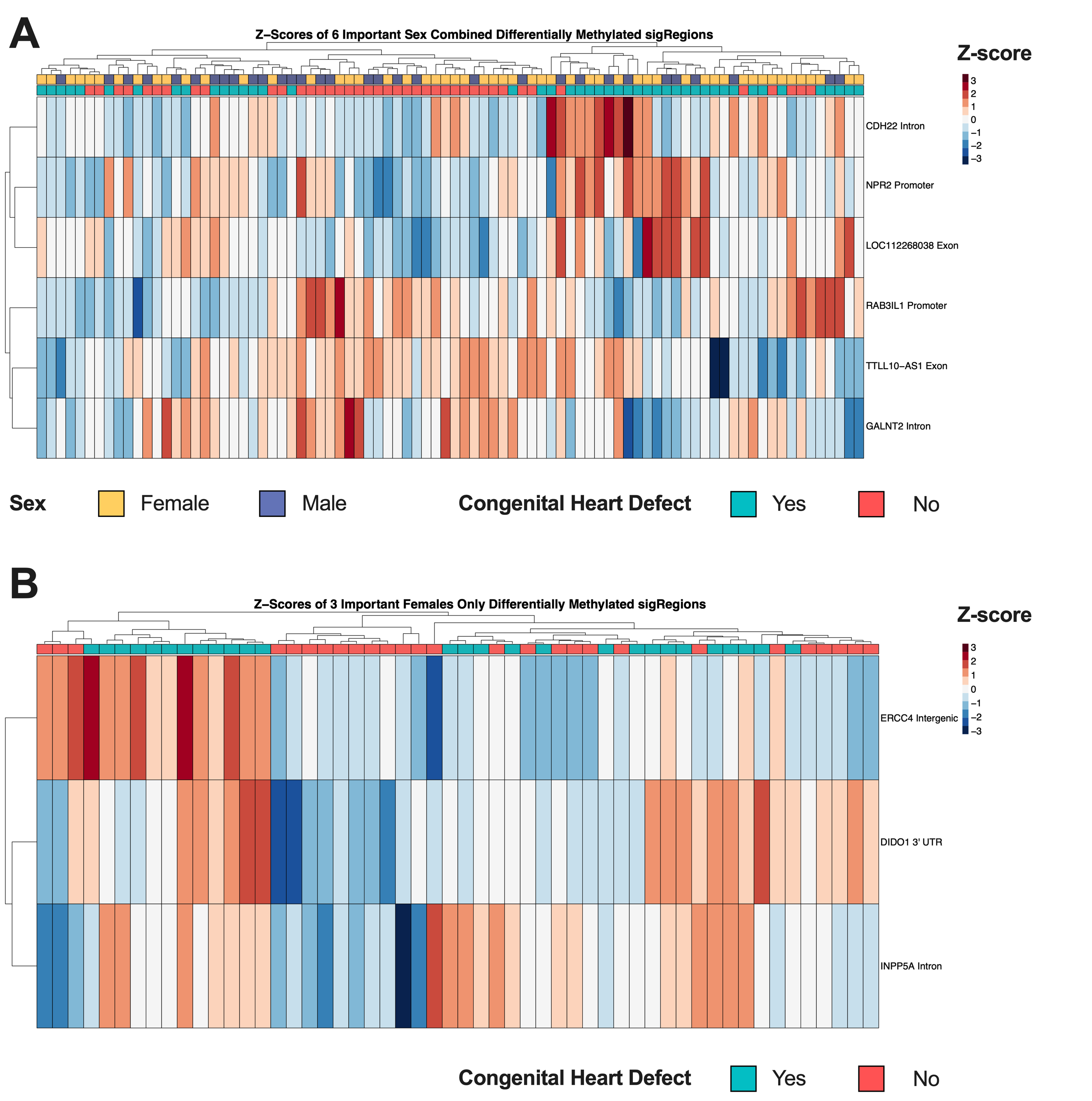

**Supplemental Figure S4.** Hierarchical clustering heatmap of the machine learning feature selection analysis of the consensus DMRs from **A)** Sex Combined and **B)** Females Only comparisons

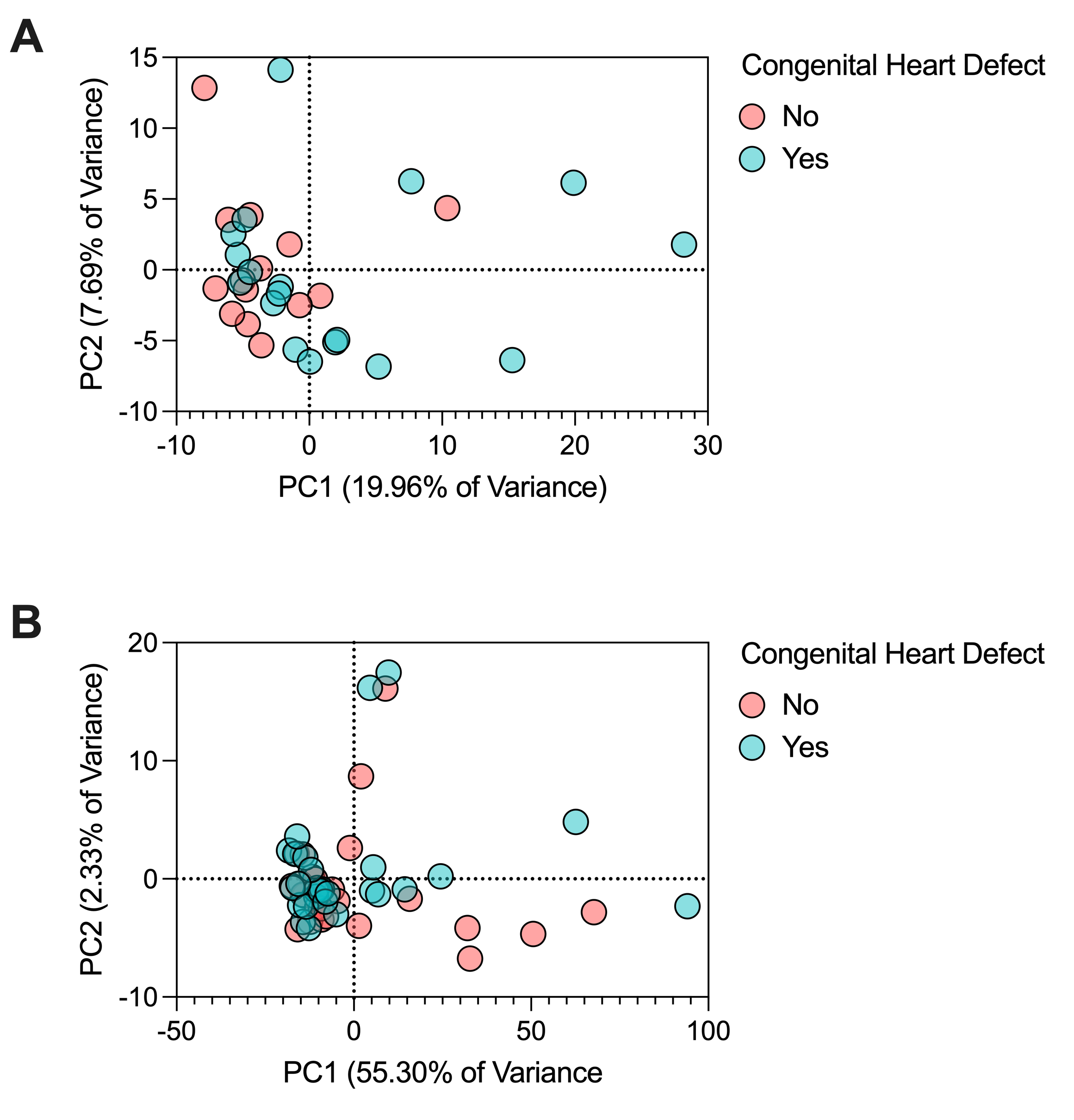

**Supplemental Figure S5.** Sex specificity of DMRs. Principal component analysis of smoothed methylation values over DMRs from the **A)** Females Only comparison in male samples and **B)** Males Only comparison in female samples. All DMRs from the Females Only comparison were used and the top 1000 most significant DMRs in male samples from the Males Only comparison were used.

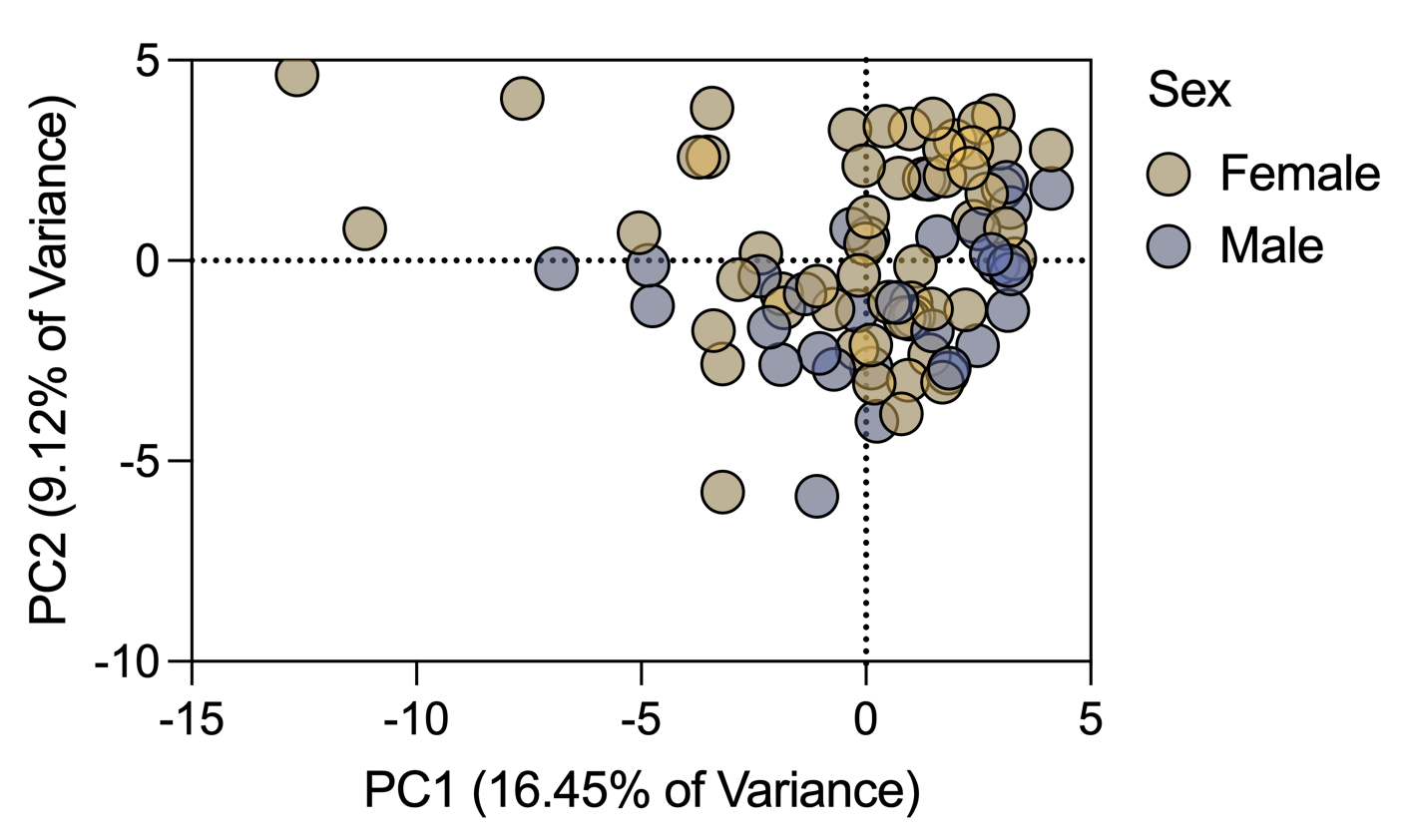

**Supplemental Figure S6.** Sex Combined DMRs do not separate by sex. PCA analysis using the smoothed methylation values of all DMRs from the Sex Combined comparison, colored by sex.

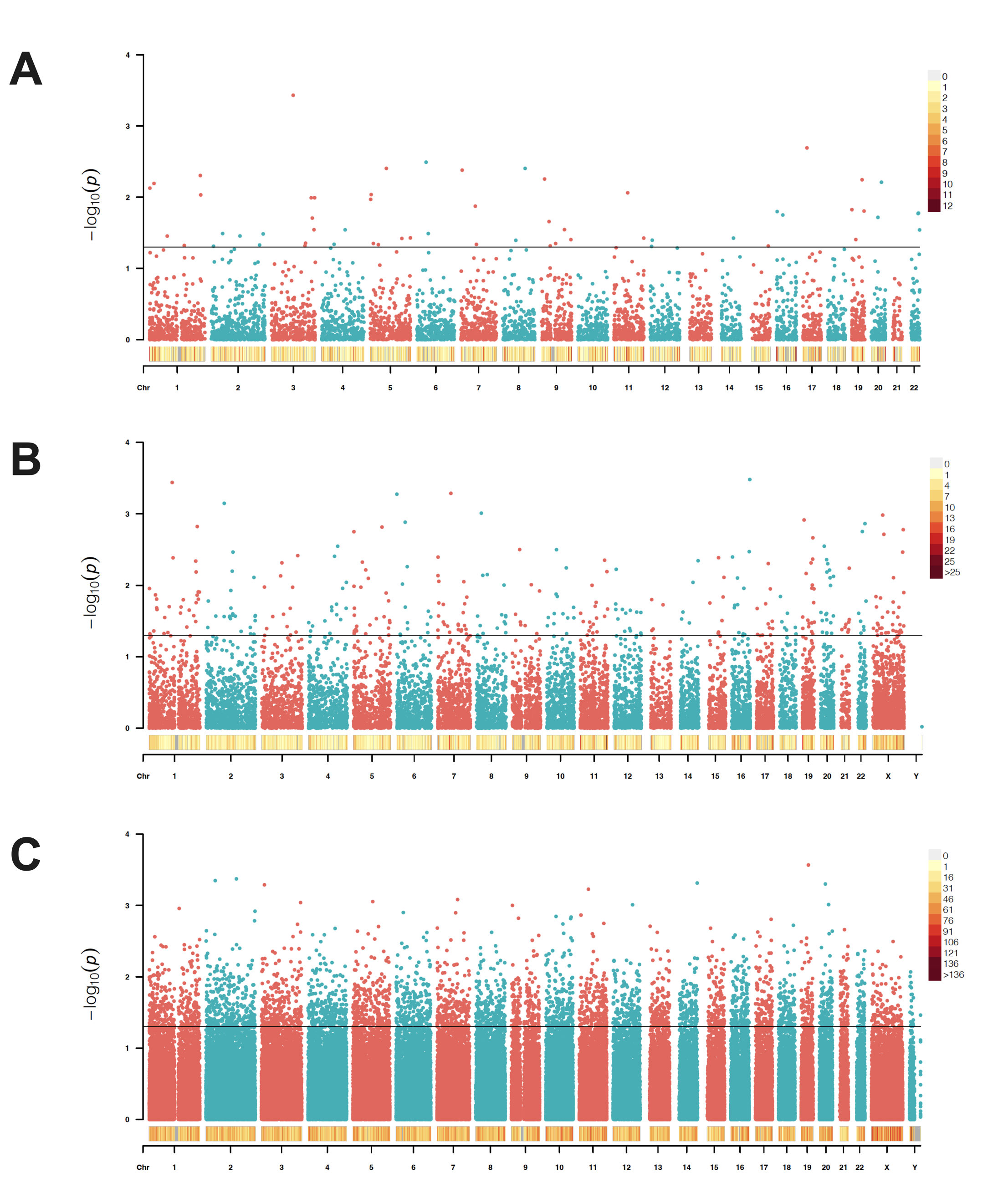

**Supplemental Figure S7**. Manhattan plots of tested background regions for the **A)** Sex-combined, **B)** Females Only, and **C)** Males Only comparisons. The density heatmap indicates the number of DMRs in 1 Mb bins and the line in the Manhattan plot indicates non-adjusted *p* = 0.05.

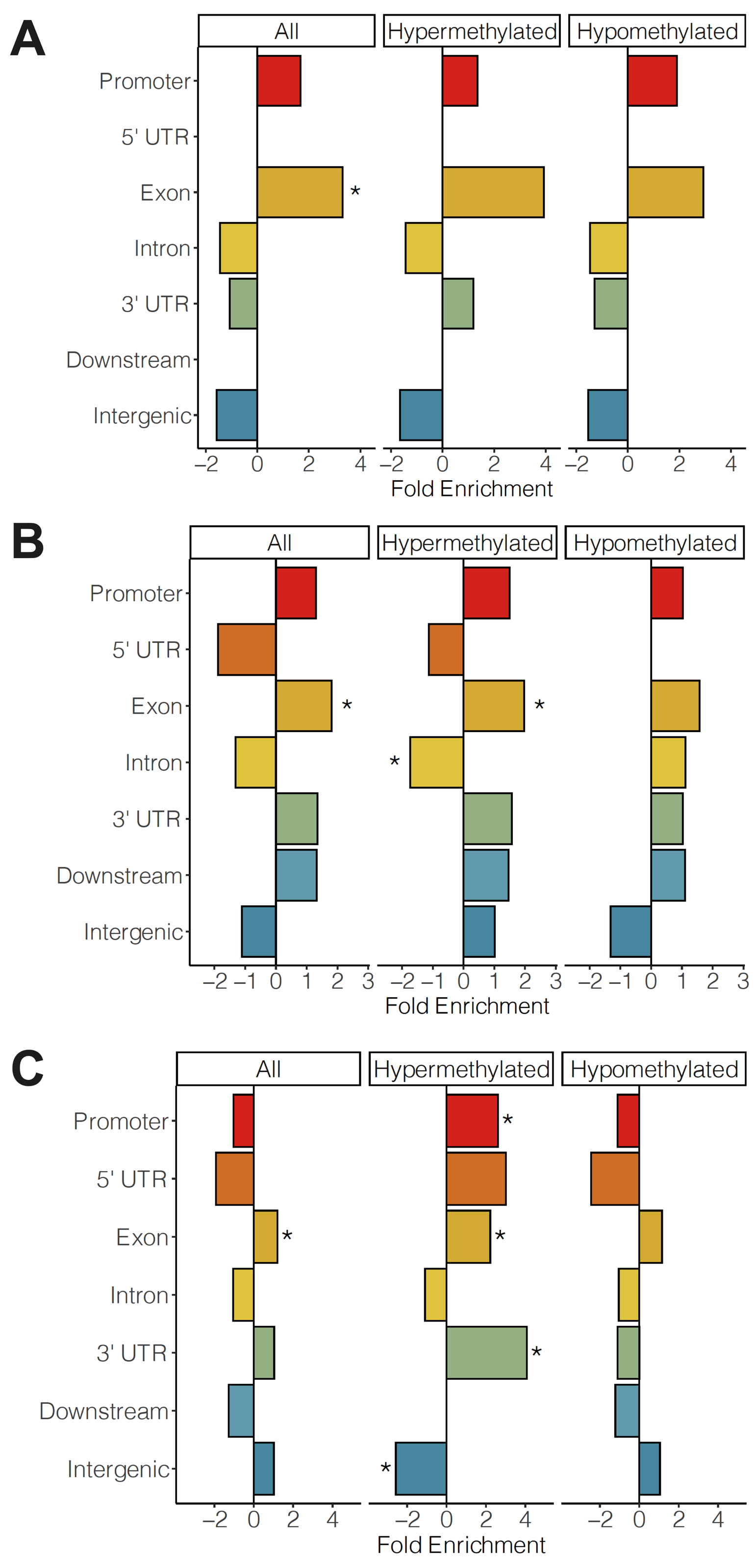

**Supplemental Figure S8.** Genic annotation enrichments of all DMRs, hypermethylated DMRs, and hypomethylated DMRs in **A)** Sex-combined, **B)** Females Only, and **C)** Males Only comparisons. * = q <0.05.

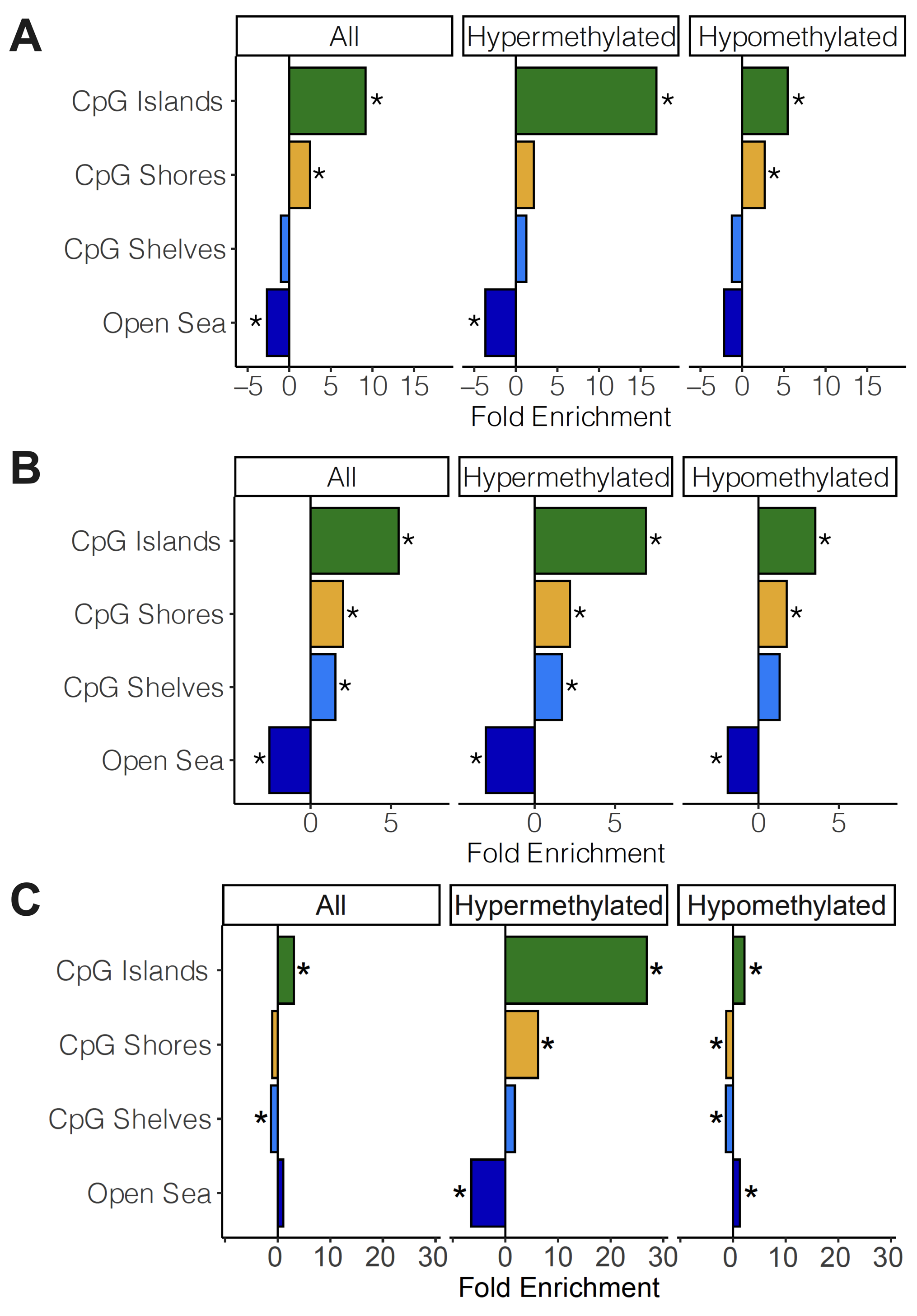

**Supplemental Figure S9.** CpG annotation enrichments of all DMRs, hypermethylated DMRs, and hypomethylated DMRs in **A)** Sex-combined, **B)** Females Only, and **C)** Males Only comparisons. * = q <0.05.

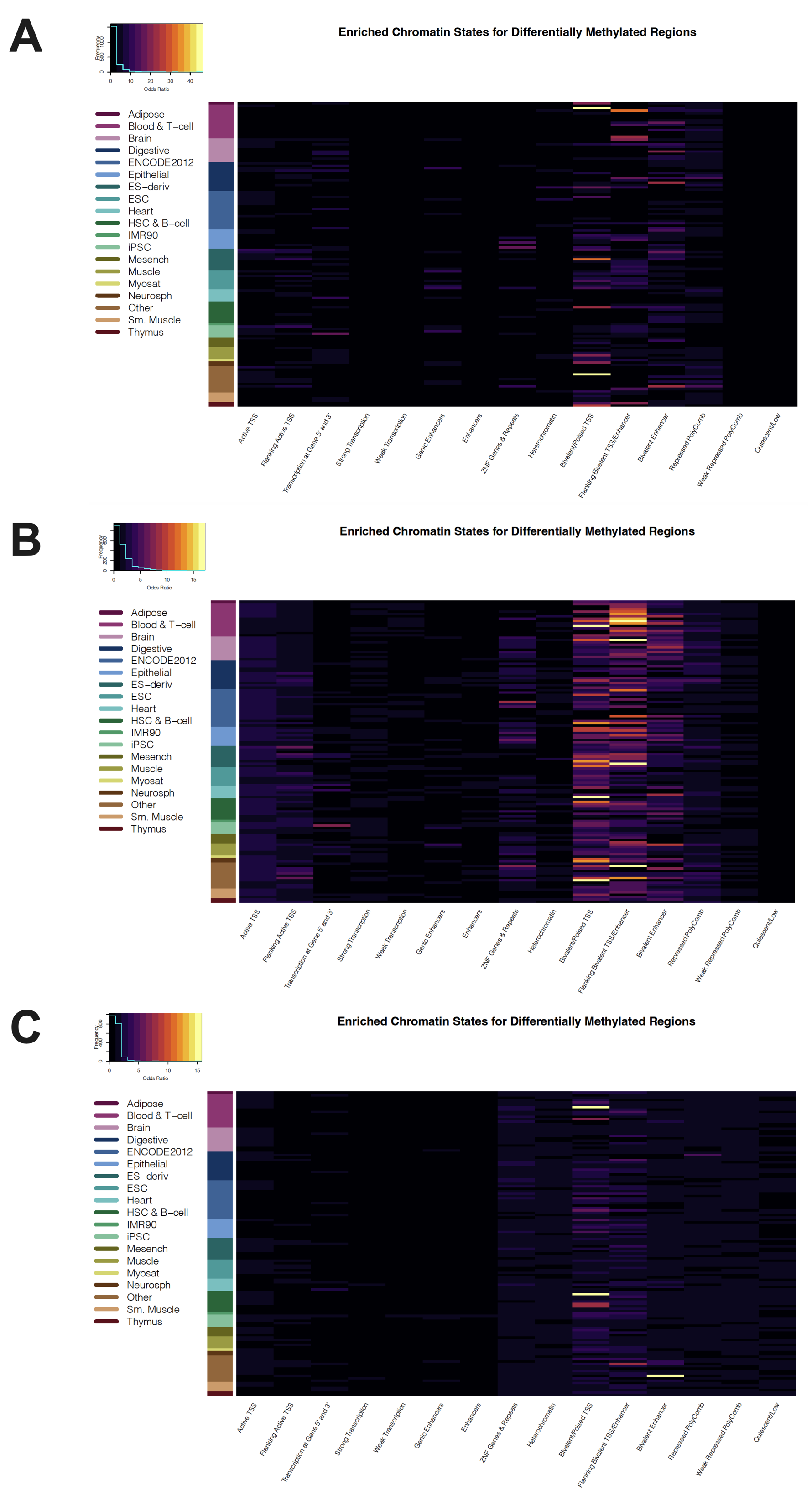

**Supplemental Figure S10.** Heatmap of odds ratios for roadmap epigenomics 127 reference epigenomes core chromatin state enrichments for all DMRs for DS CHD vs DS non-CHD in **A)** Sex-combined comparison **B)** Females Only comparison **C)** Males Only comparison

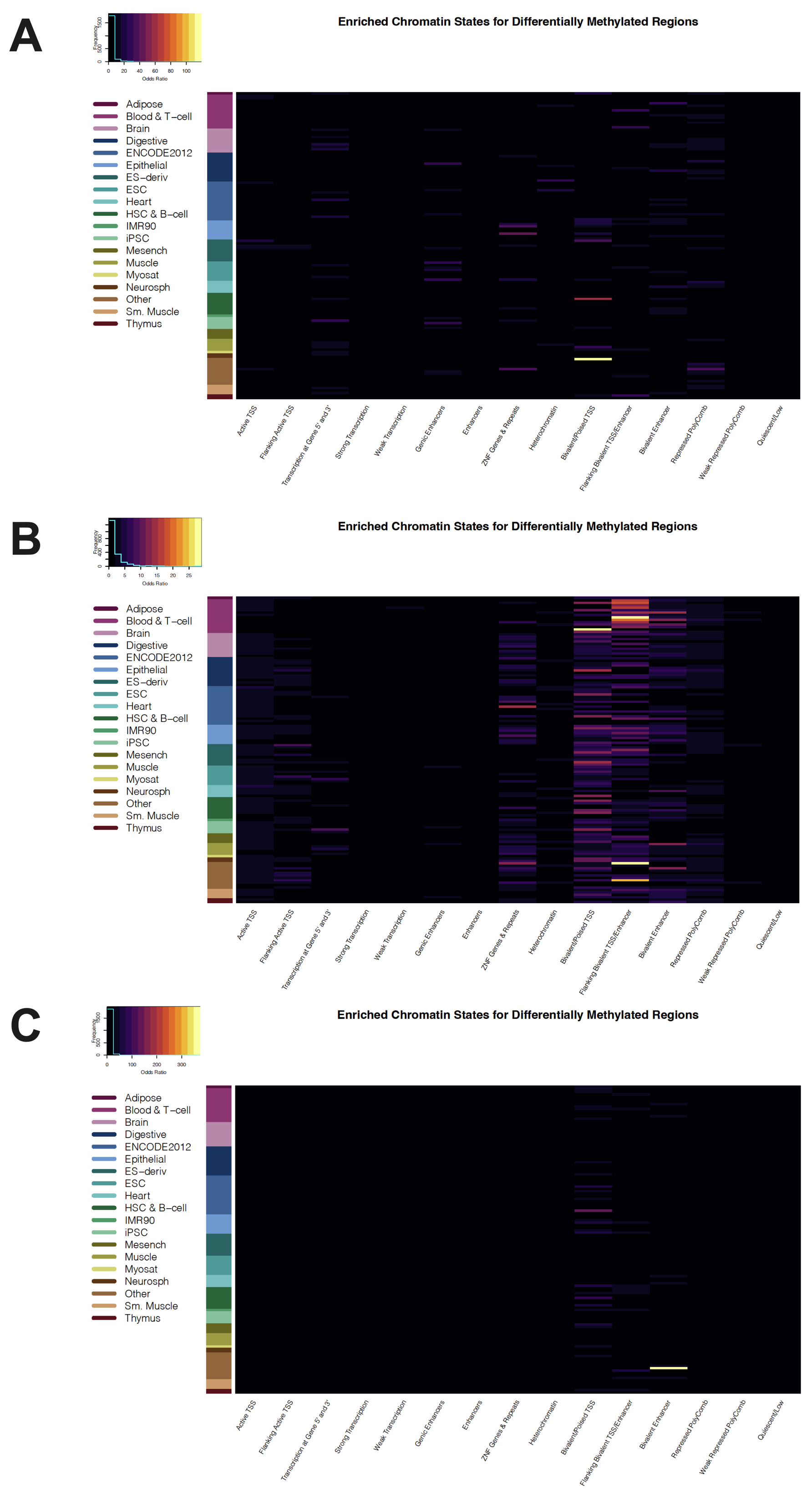

**Supplemental Figure S11**. Heatmap of odds ratios for roadmap epigenomics 127 reference epigenomes core chromatin state enrichments for hypermethylated DMRs for DS CHD vs DS non-CHD in **A)** Sex-combined comparison **B)** Females Only comparison **C)** Males Only comparison

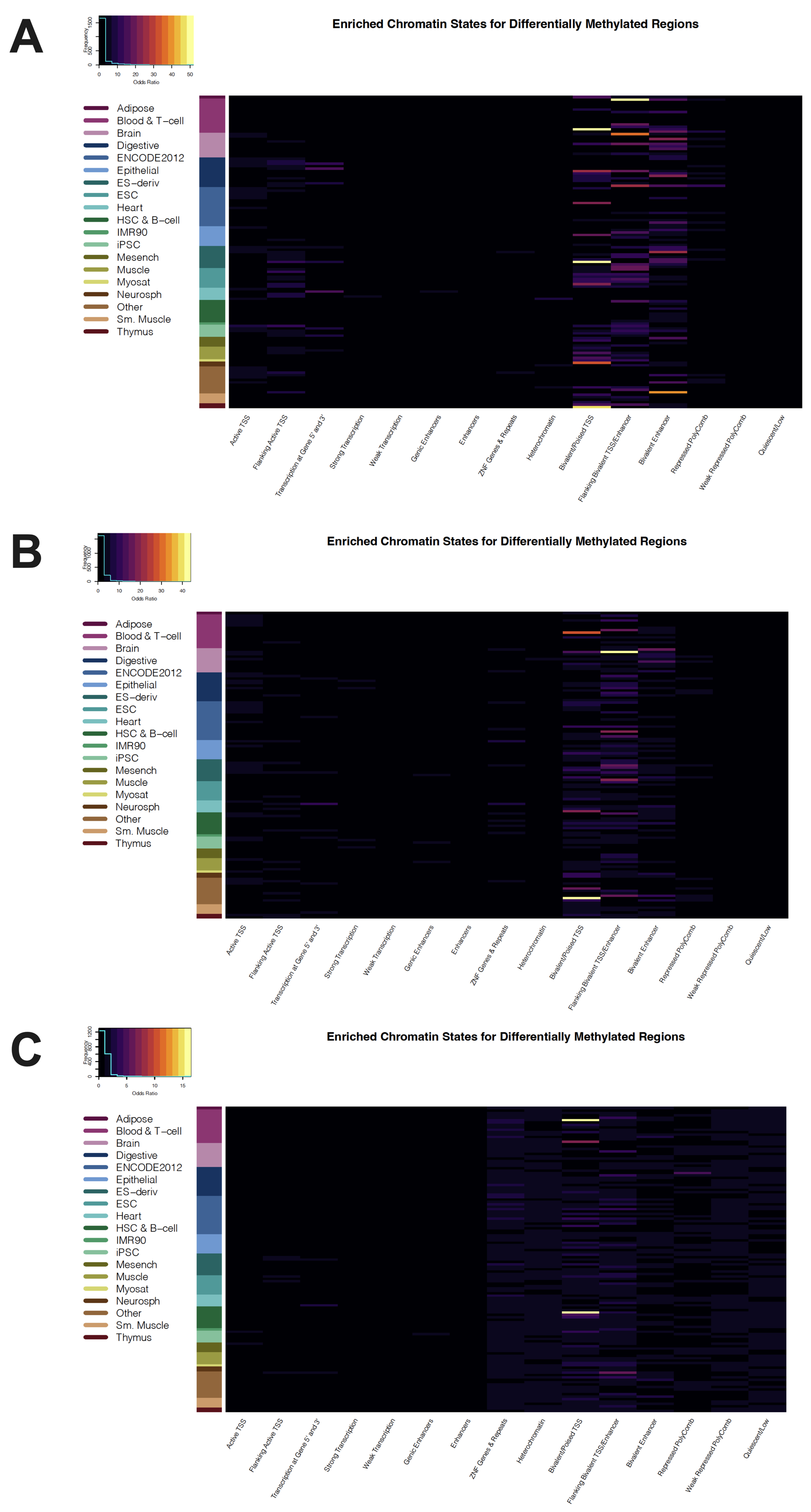

**Supplemental Figure S12**. Heatmap of odds ratios for roadmap epigenomics 127 reference epigenomes core chromatin state enrichments for hypomethylated DMRs for DS CHD vs DS non-CHD in **A)** Sex-combined comparison **B)** Females Only comparison **C)** Males Only comparison

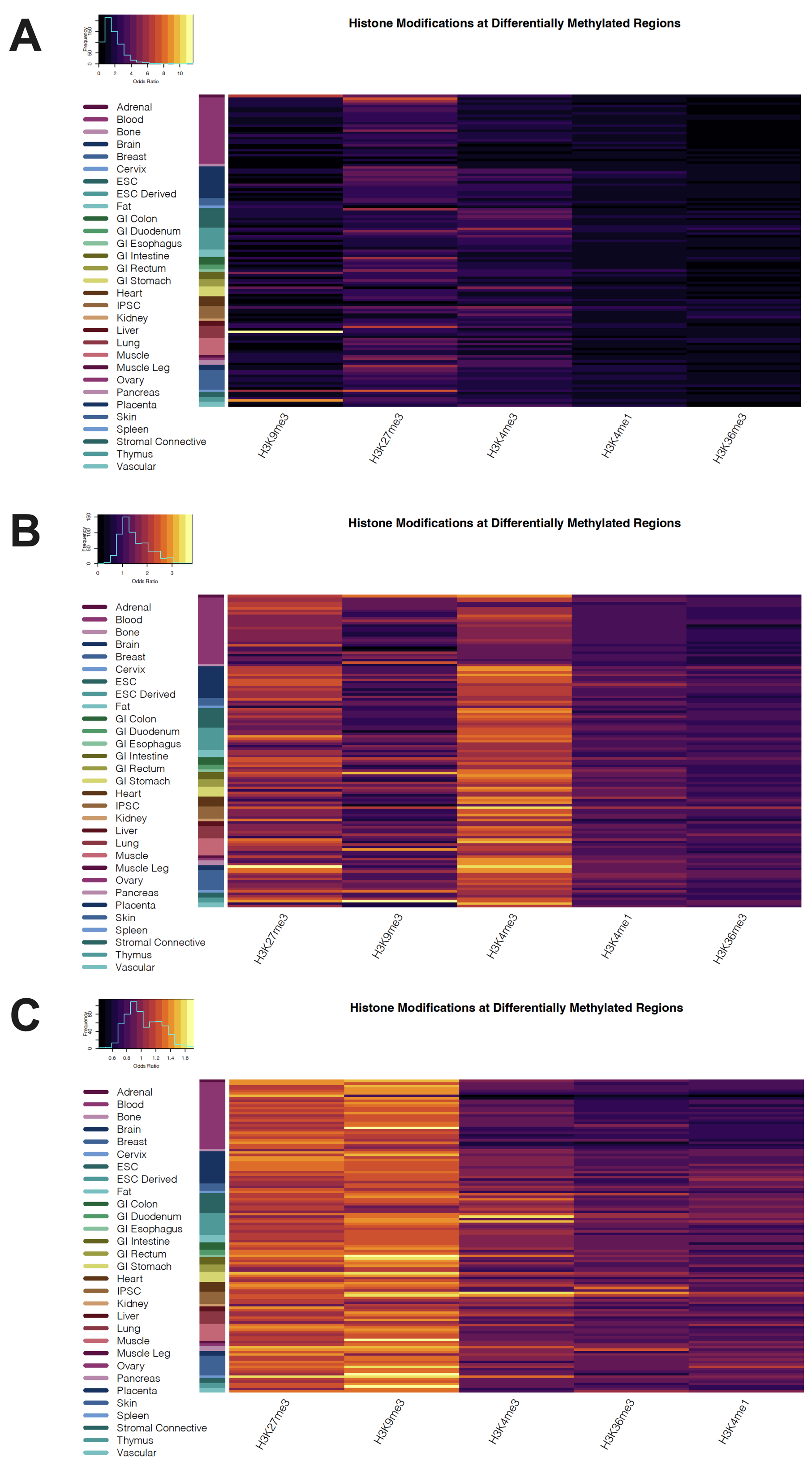

**Supplemental Figure S13**. Heatmap of odds ratios for roadmap epigenomics 127 reference epigenomes core histone modifications enrichments for all DMRs for DS CHD vs DS non-CHD in **A)** Sex-combined comparison **B)** Females Only comparison **C)** Males Only comparison

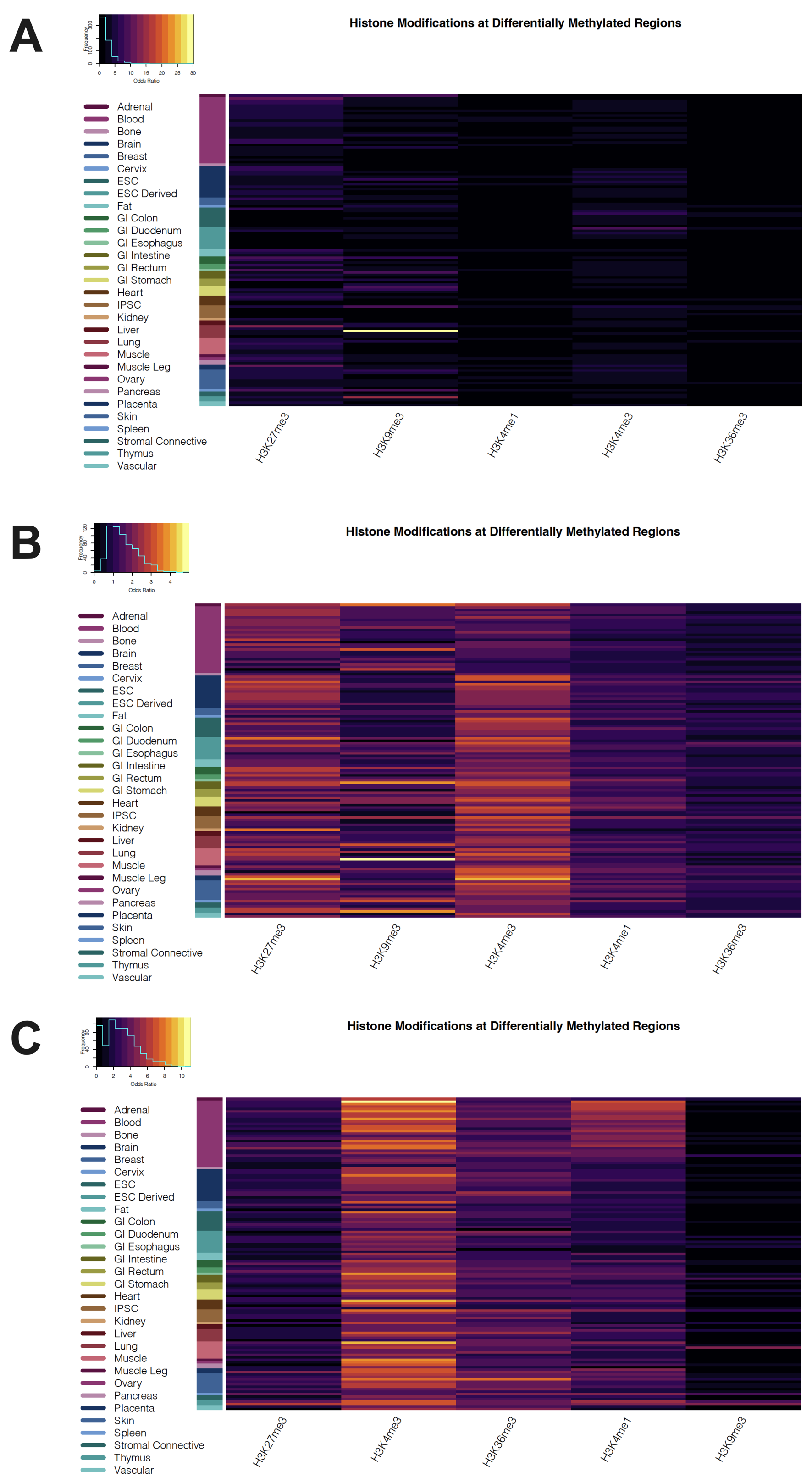

**Supplemental Figure S14**. Heatmap of odds ratios for roadmap epigenomics 127 reference epigenomes core histone modifications enrichments for hypermethylated DMRs for DS CHD vs DS non-CHD in **A)** Sex-combined comparison **B)** Females Only comparison **C)** Males Only comparison

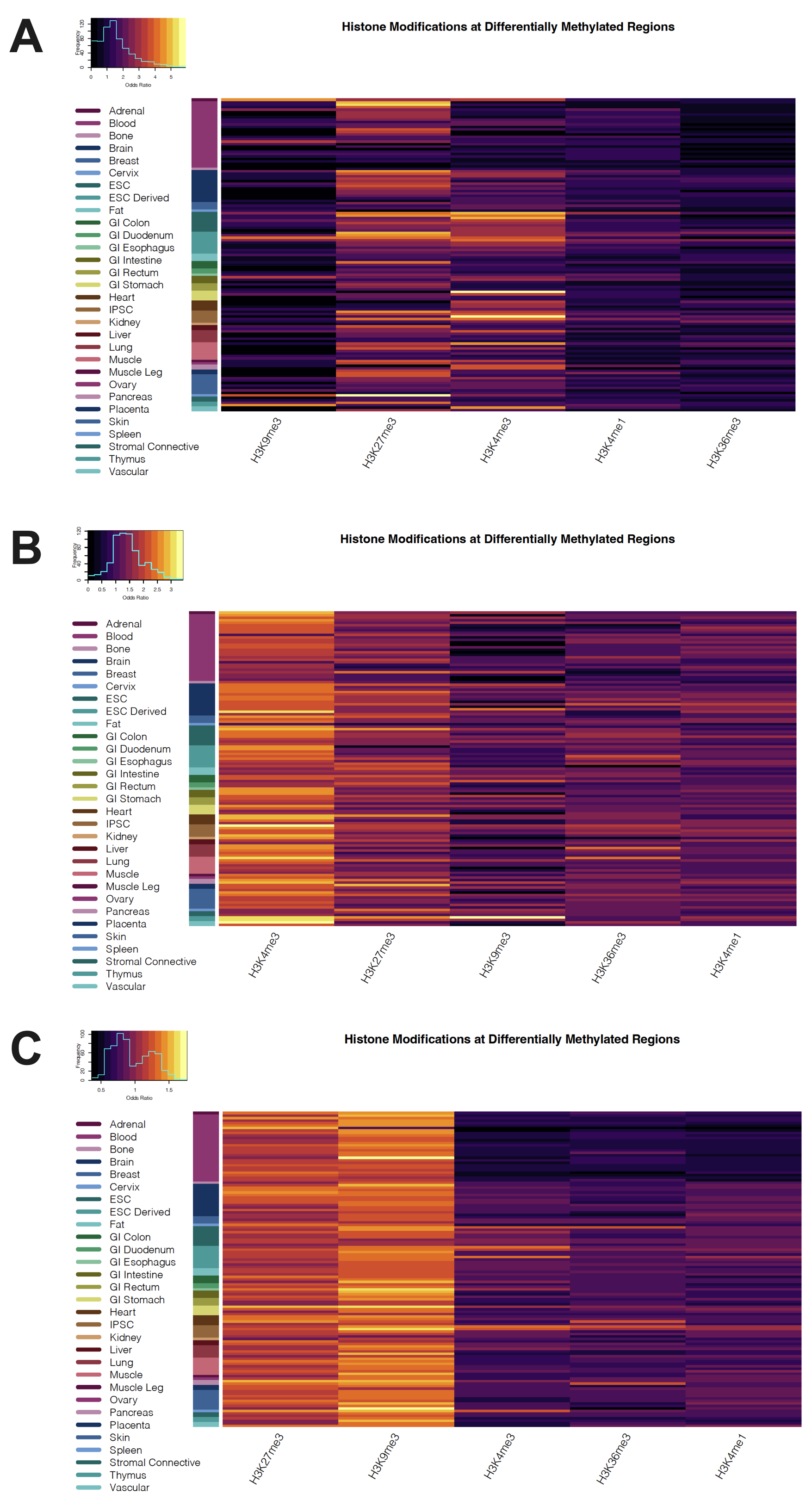

**Supplemental Figure S15**. Heatmap of odds ratios for roadmap epigenomics 127 reference epigenomes core histone modifications enrichments for hypomethylated DMRs for DS CHD vs DS non-CHD in **A)** Sex-combined comparison **B)** Females Only comparison **C)** Males Only comparison

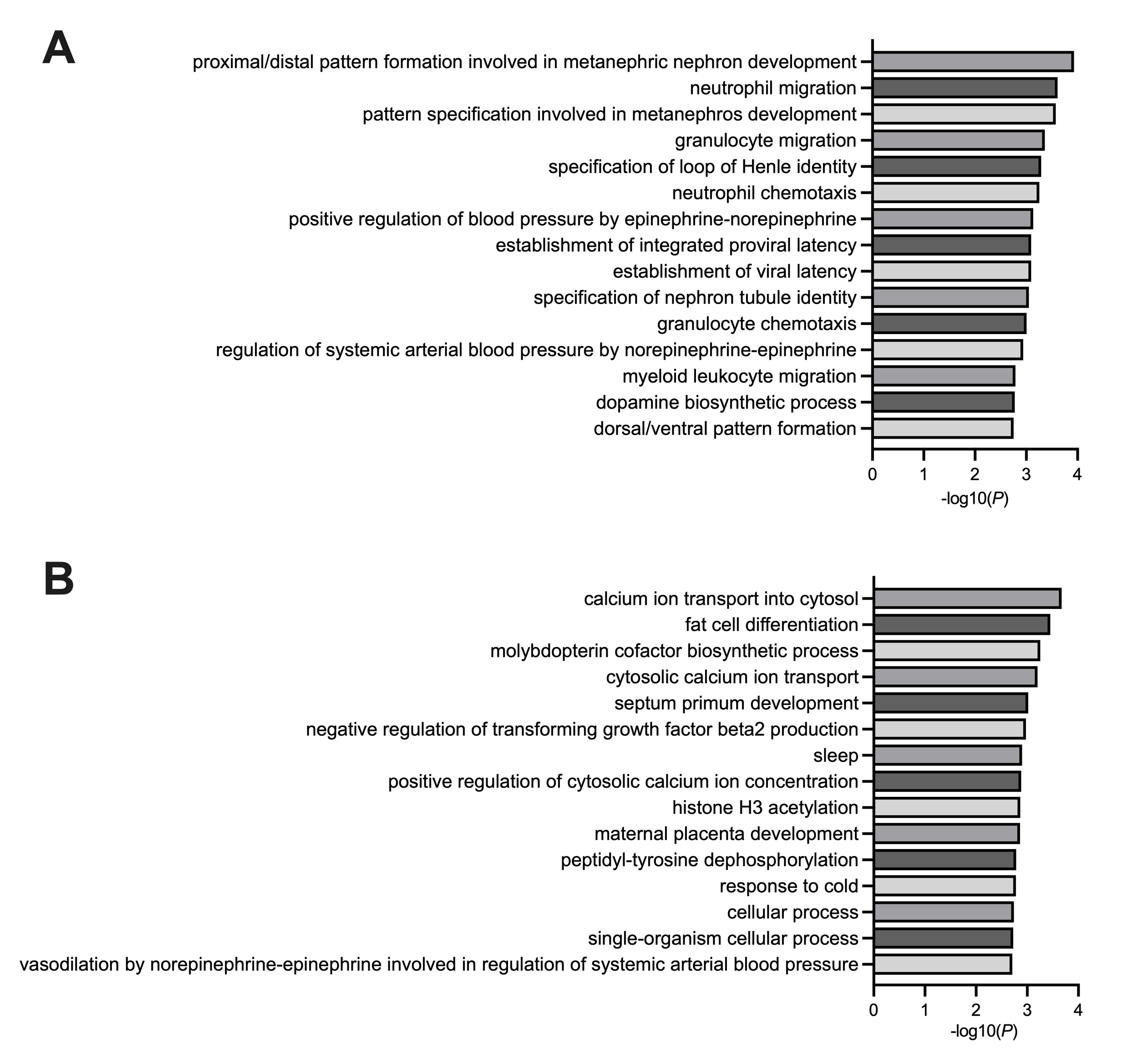

**Supplemental Figure S16.** Gene ontology enrichments. Bar plot of the fifteen most significant GO enrichments for biological processes in DS-CHD versus DS non-CHD DMRs from the **A)** Females Only comparison and **B)** Males Only comparison
